# SARS-CoV-2 neutralising antibody profiles reveal variant specific antibody dynamics and regional differences in infection histories in Malawi

**DOI:** 10.64898/2026.04.16.26351029

**Authors:** Mhairi J. McCormack, Louis Banda, Stephen Kasenda, Ellen C. Hughes, Amelia Crampin, Abena S. Amoah, Jonathan M. Read, Antonia Ho, Brian J. Willett, James A. Hay

## Abstract

Serological data provide important insights into SARS-CoV-2 transmission and immunity, particularly in regions with limited routine surveillance such as sub-Saharan Africa. However, antibody waning and boosting following reinfection or vaccination remain poorly characterised, complicating interpretation of serological measurements. Improved understanding of these dynamics is critical for accurate epidemiological inference. Modelling longitudinal serological data provides a means to quantify antibody kinetics and reconstruct infection histories. We analysed 15,679 neutralising antibody (nAb) titres from 1,675 unvaccinated, HIV-uninfected participants in urban (Lilongwe) and rural (Karonga) Malawi (February 2021 - April 2022). NAb titres against ancestral B.1, Beta, Delta, and Omicron (BA.1/BA.2) viruses were measured using an HIV-based SARS-CoV-2 pseudotyped virus neutralisation assay. A multi-level Bayesian model was used to reconstruct infection histories and antibody kinetics. The model identified 429 infections (95% credible interval 417-441), including 39 (9·1%) that had not been identified by traditional seroconversion-based thresholds. Antibody levels waned rapidly, with 48% (0·403-0·560) of the acute boost remaining after three months and only 5% (0·027-0·098) after one year. Pre-Omicron infections generated stronger antibody boosts than Omicron infections. Responses varied, with individuals clustering into low and high responders. Cross-reactive responses extended across substantial antigenic distances - Omicron infections induced broader immunity. Seroincidence was higher in Lilongwe than in Karonga (0·41 vs. 0·27 infections per person per three months), driven by the early 2022 Omicron wave. Reinfections were common, particularly among adults and urban residents. SARS-CoV-2 nAb responses following infection were heterogeneous and declined rapidly. This rapid waning underscores the importance of vaccination for sustained protection, while cross-reactivity suggests only partial immunity from prior variants. Identifying reinfections is essential for understanding transmission and finding populations at higher repeat infection risk, particularly where routine surveillance is limited.

## Introduction

Severe acute respiratory syndrome coronavirus 2 (SARS-CoV-2) has caused repeated waves of infection globally, yet the timing, magnitude, and variant composition of these waves have varied substantially between regions. In Africa, routinely reported case counts and deaths substantially underestimate SARS-CoV-2 circulation [1]. This reflects limited diagnostic testing [2] and contact tracing (particularly during 2020 and 2021), and a high proportion of asymptomatic or mild infections [3]. Population-based serological studies, in contrast, reveal extensive undetected transmission. Longitudinal evidence on infection-derived immunity remains sparse in many African populations. Meta-analyses indicate that seroprevalence in Africa rose from ∼3% in mid-2020 to over 65% by late 2021, exceeding reported case counts vastly and showing substantial heterogeneity between countries and urban versus rural settings [4].

Malawi experienced four SARS-CoV-2 epidemic waves within the first two years following emergence. These were driven by the ancestral B.1 virus from June to August 2020, the Beta variant from December 2020 to April 2021, the Delta variant from June to September 2021, and Omicron BA.1 and BA.2 from December 2021 to January 2022 [5]. Malawi was among the few African countries that did not implement a nationwide lockdown [6,7]. Previous serosurveillance in Malawi has largely focused on cumulative seroprevalence in this predominantly under-vaccinated population. Early studies reported low seropositivity at the end of 2020 (9·3% and 18·5%) [8,9], consistent with under-detection during the initial epidemic waves. By mid-2021, seroprevalence increased substantially in urban areas, reaching 81·7% in Blantyre and 71·0% in Mzuzu, with measurable neutralising activity against ancestral B.1 and Beta variants [9]. Subsequent emergence of Omicron coincided with further seroprevalence increases and rising vaccine coverage [10]. However, most studies relied on cross-sectional^10^ or convenience samples, including healthcare workers [8] and blood donors [9], limiting inference on infection timing, reinfection, and antibody kinetics.

Notably, the majority of serological studies rely on group-level titres or apply arbitrary seropositivity thresholds that do not account for time since infection, reinfection, or variant-specific responses. These challenges are amplified for SARS-CoV-2, given high reinfection rates and the emergence of successive, antigenically distinct variants [11]. Modelling frameworks that integrate antibody kinetics with exposure history and variant-specific responses are therefore essential for extracting meaningful inferences from longitudinal data.

An additional complexity arises from the difficulty of distinguishing variant-specific responses from cross-reactive antibodies induced by antigenically similar variant. Antigenic cartography offers a framework to disentangle these relationships, using neutralisation data to quantify antigenic distances and infer cross-reactivity [12-15]. This can be extended through antibody landscapes extend this framework to capture both the magnitude and breadth of responses across antigenic space over time. Mechanistic serodynamic models offer an additional complementary approach, explicitly modelling antibody kinetics, cross-reactivity, reinfection, and measurement error. Mathematical models have been applied to estimate real-time immune responses to emerging SARS-CoV-2 variants in the UK, quantifying variant-specific neutralisation and waning dynamics [16]. However, these approaches have rarely been evaluated in African populations, where repeated natural infection dominates and longitudinal data are sparse. *Serosolver* [17], a modelling framework originally developed for influenza [18,19], can reconstruct individual infection histories from longitudinal antibody data, but it has not yet been used to jointly infer infection histories alongside multi-variant nAb dynamics after repeated SARS-CoV-2 infection.

Longitudinal characterisation of antibody trajectories in Africa remain limited, and existing studies have not incorporated nAb data [20-22]. This represents a critical gap, as nAb titres provide a functional correlate of protection against infection and severe disease [23], and their longitudinal dynamics offer insight into the development, waning, and breadth of immunity in populations experiencing repeated natural exposures. Existing studies have also been limited by incomplete quantification of antibody waning; lack of coverage of Omicron-dominant waves; and no incorporation of individual-level immune heterogeneity.

In this study, we analysed nAb data from unvaccinated, HIV-uninfected individuals in urban and rural Malawi, collected between February 2021 and April 2022 as part of the COVSERO study [5,24]. Previous analyses characterised IgG [5] and nAb [24] responses using group-level comparisons, assessing overall positivity, and response magnitude across demographic and geographic strata. Here, we applied the *serosolver* R package to reconstruct individual infection histories, assess the strength and durability of neutralising responses across multiple variants, and explore heterogeneity in exposure and immunity across subgroups. Our aim was to reconstruct the epidemiological history of SARS-CoV-2 in this population, characterising seroincidence, while examining the kinetics and persistence of nAb responses and accounting for the complexities of nAb analyses. By integrating continuous neutralisation titre data, antigenic distances, and robust infection history modelling, we provide a nuanced understanding of transmission and immunity in a low-surveillance, low-vaccination setting and offer insight into how successive epidemic waves shape population-level immunity.

## Materials and Methods

### Ethics statement

Ethical approval for this study was obtained from the Malawi College of Medicine Research Ethics Committee (P11/20/3177, approved on 11 December 2020) and the University of Glasgow College of Medicine, Veterinary and Life Sciences Research Ethics Committee (200200056, approved on 8 February 2021). All study procedures complied with applicable laws and institutional regulations, and the research was conducted in accordance with the principles of the Declaration of Helsinki. Written, informed consent was obtained from participants or guardians for minors and vulnerable adults.

### Study design

We conducted a modelling study using data from a population-based longitudinal study, with samples collected from 24^th^ February 2021 to 22^nd^ April 2022 in urban and rural communities in Malawi.

### Study cohort and sample collection

The cohort included individuals over one year old from randomly selected households within the Malawi Epidemiology Intervention and Research Unit (MEIRU) population cohorts (n=2,005), recruited from a rural site (Karonga Health and Demographic Surveillance Site (HDSS)) and an urban site (Area 25, Lilongwe) (S1 Fig) [5]. Participants were enrolled between 24 February and 8 June 2021 (Survey 1) and followed at three-month intervals across three additional surveys: Survey 2 (28 June–13 September 2021), Survey 3 (4 October–10 December 2021), and Survey 4 (27 January–22 April 2022) (S2 Fig). The participant selection process and sample size calculation was described in Banda et al. 2023 [5]. Only COVID-19 unvaccinated, self-reported HIV-uninfected individuals with sufficient serum were included (n=1,675), and participants were removed once vaccinated to capture natural infections only (number removed per survey – Survey 1: n=94; Survey 2: n=195; Survey 3: n=239; Survey 4: n=267).

Baseline data were collected via interviewer-administered questionnaires, covering demographics, socioeconomic status, medical history, COVID-19 symptoms, vaccination, and prevention practices. Follow-up surveys captured updated vaccination status, symptoms, and healthcare use. Venous blood samples were collected at each round, serum was stored at –80⍰°C at the MEIRU biobank, and later transported to the Medical Research Council (MRC)–University of Glasgow Centre for Virus Research for laboratory analyses. Participant age per study site was presented as median with interquartile range (IQR) and compared using the non-parametric Wilcoxon rank-sum test.

### SARS-CoV-2 serology

SARS-CoV-2 neutralising activity was measured using an HIV(SARS-CoV-2) pseudotyped virus neutralisation assay (PVNA), with cell lines and pseudotype generation methods described in the S1 Supplementary Information. All samples were initially screened at a single dilution against ancestral B.1, Beta, Delta, and Omicron BA.1 (S1 Table). Samples showing >90% neutralisation against any variant were classified as positive and subsequently titrated against variants circulating in Malawi at each survey: Survey 1 – ancestral B.1, Alpha, Beta, Delta; Survey 2 – ancestral B.1, Beta, Delta, Omicron BA.1; Survey 3 – ancestral B.1, Beta, Delta, Omicron BA.1; Survey 4 – ancestral B.1, Beta, Delta, Omicron BA.1, Omicron BA.2 (S1 Table). Neutralising titres were defined as the serum dilution achieving 90% reduction in infectivity.

### Model overview

RStudio version 2024.12.0+467 was used for mathematical modelling and data interpretation.

We applied *serosolver* [17], an open-source R package that infers antibody kinetics, individual infection histories, and population-level SARS-CoV-2 seroincidence from longitudinal nAb data. The mathematical model describes transmission with time-varying infection rates and antigenic evolution of circulating variants (S3 Fig). Each infection is assumed to elicit antibody boosting followed by waning, and cross-reactive responses are determined by the antigenic distance between the infecting variant and the measured variant.

Model inputs include longitudinal nAb data, demographic variables, an antigenic map of SARS-CoV-2 variants, and priors on all parameters. Antibody data comprised participant ID, sample timing (3-month intervals, Q1 2021–Q2 2022), assay variant, and log_3_(titre/50) titres. The model accounted for exposures beginning in Q1 2020. Demographic variables included site (urban/rural) and age (<15 years, n=605; ≥15 years, n=1,070). Exploratory analysis indicated that, for a subset of individuals, nAb responses were substantially higher than predicted by the population-level model, suggesting that boosting was underestimated and that a single response profile was insufficient. To better capture this heterogeneity, individuals were classified as either high responders (titre >300, log_3_>1·63; n=29) or low responders (n=1,646). As no standardised cutoff defines “high” versus “low” responders, this threshold was selected empirically based on the distribution of titres in our cohort to distinguish individuals with markedly stronger neutralising responses from the broader population. Antigenic distances were calculated as Euclidean distances between variant coordinates on an antigenic map [15], and weakly informative priors were applied to all parameters (S2 Table).

*Serosolver* uses Markov chain Monte Carlo (MCMC) to estimate posterior distributions for parameters describing boosting, waning, and cross-reactivity, enabling population-level summaries and comparisons. It predicts individual antibody titres over time, with observed versus predicted values visualised to assess fit. It also infers posterior infection probabilities, allowing estimation of seroincidence and reinfection patterns. Individual infection events were classified as infections when the median posterior probability of infection exceeded 0·5, indicating that infection was more likely than not under the model. For reinfections, events were linked to demographic factors, and age-, residence-, and response-specific risk ratios were calculated using 2×2 contingency tables and the *riskratio* function from the *epitools* R package (v0.5-10.1) [25], using a significance threshold if p<0.05. Exact p-values are displayed if p ≥ 0.001, otherwise, p < 0.001 is presented.

The core *serosolver* framework is described in detail elsewhere [17]. For this study, we implemented multiple extensions to improve model performance for SARS-CoV-2 nAb data, including adjustments to the observation and antibody kinetics components (S4 Fig, S1 Supplementary Information).

### Antibody kinetics inference

The *serosolver* model estimates key parameters describing antibody dynamics, including short-term boosting, waning, and cross-reactivity. Unlike earlier influenza applications that assumed linear waning [18], we adopted an exponential waning model on the log scale [26], which ensures predicted titres for previously infected individuals remain detectable despite waning (S4 Fig).

The complete antibody kinetics model is expressed as *f*(*A*|*Z, θ*), with *A* denoting true antibody levels, *z* infection histories, and *θ* = {*μ*_*s*_, *μ*_*l*_, *ω, σ*_*l*_, *σ*_*s*_, *τ*}. For individual *i*, infected at times *j* and sampled at *t*, the predicted antibody titre is:

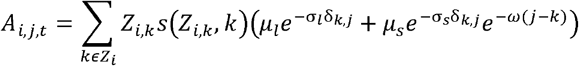

Thus, each observed antibody titre reflects the cumulative effect of all prior infections, combining a rapidly waning short-term response with a long-lasting boost, modulated by antigenic distance and the sequence of prior exposures. Exploratory analysis suggested that long-term boosting, *µ*_*l*_, could not be identified alongside short-term boosting, *µ*_*S*_, and thus we fixed *µ*_*l*_ = 0.

### Stratifications on antibody kinetics

In the original *serosolver* framework [17,18], all individuals shared the same antibody kinetics parameters across infections with different strains or variants. Here, short-term boosting (*µ*_*S*_) and short-term cross-reactivity (*σ*_*S*_) were stratified by response type (S2 Table), capturing variation between groups of individuals. We also stratified short-term boosting, cross-reactivity, and waning by pre-Omicron and Omicron exposures, reflecting evidence that Omicron infections generally generate weaker boosting and altered cross-neutralisation, effectively behaving as a distinct serotype [27]. Infections during the Omicron period were governed by Omicron-specific parameters, while pre-Omicron infections used pre-Omicron parameters. The model assumed no co-circulation of variants; each three-month window was assigned a single circulating variant, ensuring all infections were unambiguously linked to the corresponding variant-specific parameter set.

### Antigenic map

Since cross-reactivity was modelled as a function of antigenic distance, selection of antigenic map a critical choice. Antigenic maps vary by serum source, assay type, laboratory conditions, cartography method, and biological variability. We used the map from Wilks et al. 2023 [15], extracting coordinates for ancestral B.1, Beta, Delta, Omicron BA.1, and Omicron BA.2 (Fig 1). This map was based on 207 sera obtained from vaccinated or SARS-CoV-2-infected individuals across multiple global regions, including Africa. Infections were assumed to have had a single infection at sampling, with variants confirmed by whole-genome sequencing. All samples were tested against 21 SARS-CoV-2 variants using an HIV-based neutralisation assay. Participants living with HIV were included. However, samples exhibiting unexpected reactivity in the absence of documented vaccination or infection were excluded to avoid interference from antiretroviral therapy.

**Fig 1.**
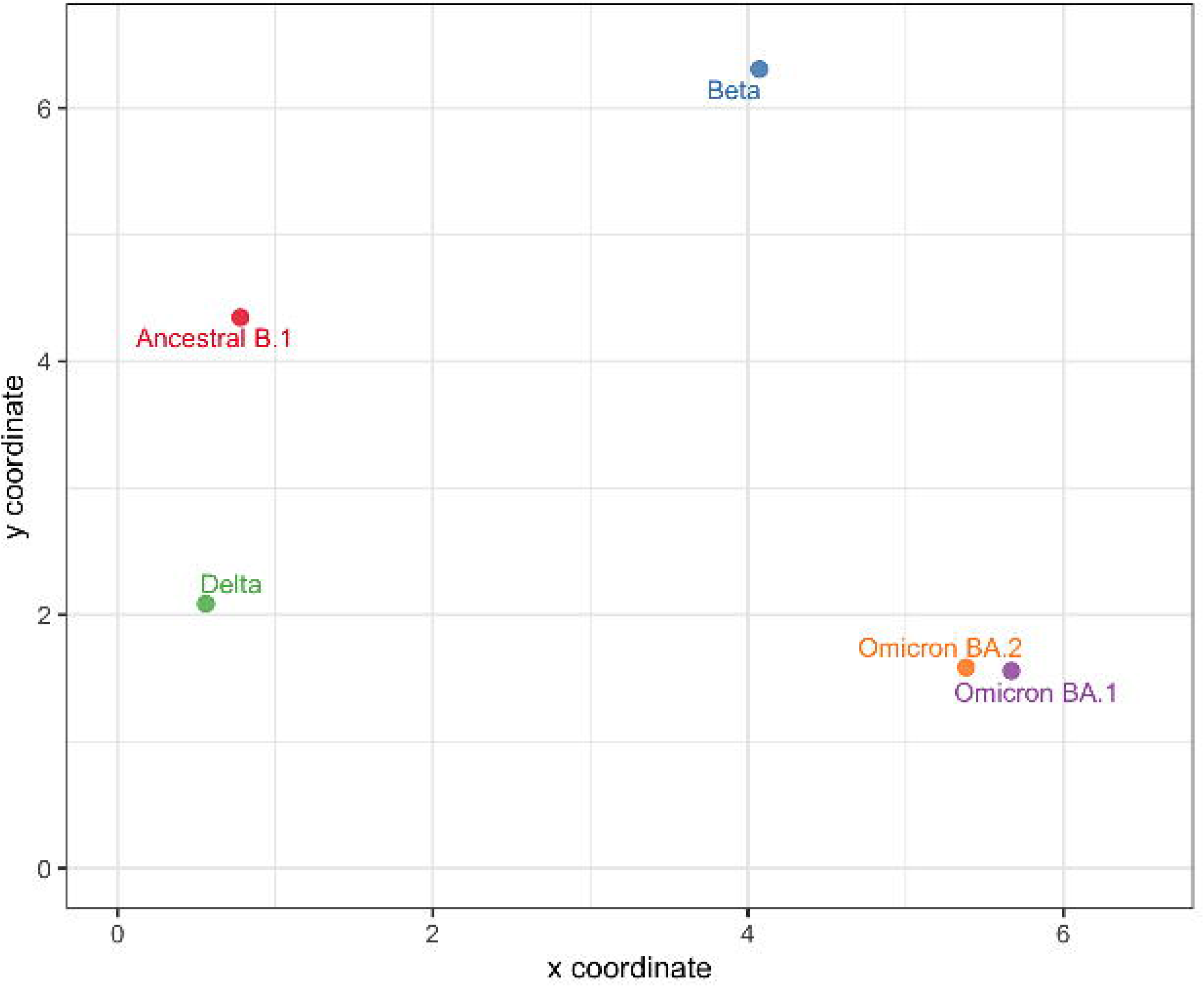
Antigenic map used in the model. Antigenic distances between SARS-CoV-2 variants are shown: ancestral B.1 (red); Beta (blue); Delta (green); Omicron BA.1 (purple); and Omicron BA.2 (orange). Map coordinates were derived from Wilks et al. 2023 [15].

Additionally, we performed sensitivity analysis, altering the antigenic map (S5 Fig) to observe any differences in model output (as described in S1 Supplementary Information

### Infection model stratification

The infection model is detailed in the S1 Supplementary Information and S6 Fig. To account for differences in exposure risk, priors on infection probabilities were stratified by study site (urban vs. rural), reflecting higher expected exposure frequency in urban settings. Within the Beta–Bernoulli framework, all participants at a given site shared a common prior on infection probability for each time window, while urban and rural cohorts retained independent priors. Thus, infection priors for urban participants follow a site-specific Beta– Binomial distribution, with a separate distribution for rural participants.

### MCMC settings

*Serosolver* estimates posterior distributions of antibody kinetics parameters and latent infection histories using a Markov chain Monte Carlo (MCMC) method. Parameter updates employ a custom Metropolis-Hastings algorithm with adaptive univariate normal proposals for continuous parameters and a specialised sampler for discrete infection states. We ran three independent chains, each with 500,000 adaptive iterations for tuning, followed by 1,000,000 sampling iterations, discarding the first 500,000 as burn-in to mitigate the influence of starting values.

## Results

### Cohort characteristics

We analysed nAb titres against five SARS-CoV-2 variants (specifically ancestral B.1 virus, and the Beta, Delta, Omicron BA.1 and Omicron BA.2 variants) in 1,675 unvaccinated participants from two regions of Malawi: Karonga (n=897, rural) and Lilongwe (n=778, urban). Serum samples were collected longitudinally at four visits between February 2021 and April 2022. To ensure accurate estimation of infection-induced antibody responses, vaccinated individuals were not included in the analysis, since the *serosolver* model cannot distinguish vaccine-induced from infection-induced antibody responses (total removed per survey - Survey 1: n=94; Survey 2: n=195; Survey 3: n=239; Survey 4: n=267). Surveys were spaced a median of 106 days apart (IQR 98–118; range 64–164) and conducted sequentially without overlap, with sampling evenly distributed within each round (S2 Fig). At the final survey, participant ages ranged from 1 to 87 years. Median age was slightly higher in the urban region (24·5 years, IQR 14·0–37·8) than in the rural region (20·8 years, IQR 11·8–38·0; p=0·024, Wilcoxon rank-sum test).

### Overview of neutralisation data

The dataset comprised 15,679 nAb measurements from four survey rounds, covering five SARS-CoV-2 variants, enabling assessment of both population-level patterns and individual-level dynamics of immune responses. Titres were positively skewed, showed clear temporal trends (including pronounced Omicron responses in Survey 4), and differed by study site (S7 Fig). Median titres were generally lower in the rural region (Karonga) than in the urban region (Lilongwe). To illustrate within-host variation, representative longitudinal titre trajectories from three individuals illustrated the heterogeneity of immune responses (S8 Fig): one with an early infection (participant 262), one with a late infection (participant 1,532), and one with a complex pattern likely representing an early infection and then reinfection (participant 936). These examples reflect variation in peak titres, antibody durability, and boosting from subsequent exposures.

### Model diagnostics overview

Convergence was assessed using both visual and quantitative diagnostics. Trace and density plots confirmed well-mixed, stationary chains, with effective sample size (ESS) greater than 200 and 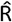 (potential scale reduction factor) below 1·1. Trace and density plots indicated unimodal posterior distributions (S9, S10 Figs). Histograms demonstrated model predictions closely matched observed titres, including negatives (S11 Fig), and point-range plots showed alignment across the full measurement range (S12 Fig), with 99·5% of observations within 95% prediction intervals. All parameters had ESS >200 and 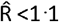, with 7/9 <1·01.

### Antibody kinetics parameter estimates

Model fitting to observed antibody trajectories estimated the short-term nAb boost posterior median estimate at 0·859 (CrI: 0·690–1·04) on the log_3_(titre/50) scale, corresponding to a titre of 128·4 (CrI: 106·7–156·9) (Table 1). The antibody boost waned rapidly (waning rate estimate: 0·734 (CrI: 0·580–0·908)), corresponding to 48% (CrI: 0·403– 0·560) of the response remaining after three months and 5% (CrI: 0·027–0·098) after one year. Because early infections were infrequent, we did not have sufficient data to estimate longer-term boosting and waning. Hence, we focused on short-term dynamics. Predicted antibody trajectories showed a steep decay, with homologous variant antibodies falling to ∼1% of the initial boost by ∼540 days and to 0·1% by ∼845 days (S13 Fig).

**Table 1.** Estimated values for the antibody kinetics parameters at baseline (low responders, infected pre-Omicron), inferred using *serosolver*. The estimated value is on the log_3_(titre/50) scale.

| Parameter | Description | Units | Estimate (posterior median; 95% CrI) – $\log_3(\text{titre}/50)$ |
| --- | --- | --- | --- |
| $\mu_s$ | Short-term boosting | $\log_3(\text{titre}/50)$ units | 0.859 (0.690–1.04) |
| $\sigma_s$ | Short-term cross reactivity | Proportion decrease in boost per unit of antigenic distance | 0.155 (0.0961–0.222) |
| $\omega$ | Waning rate parameter for the short-term response | $\log_3(\text{titre}/50)$ units per three months | 0.734 (0.580–0.908) |
| $\epsilon$ | Standard deviation of observations | $\log_3(\text{titre}/50)$ units | 0.848 (0.812–0.884) |

CrI – credible interval.

The cross-reactivity rate was 0·155 (95% CrI: 0·096–0·222), reflecting the exponential decline of nAb titres with increasing antigenic distance (Table 1). Higher values for this parameter indicate weaker cross-neutralisation against more distant variants. Using the antigenic map from Wilks et al. 2023 (15), ancestral B.1 to Omicron BA.1 distance was 5·63 units, corresponding to ∼42% retention of the homologous B.1 boost in our model. For closer variants such as Delta (distance 2·27), cross-reactivity increased to ∼70%. We applied these estimates to Omicron BA.4, subsequent variant detected in Malawi [28]. Antigenic distances between BA.1–BA.4 and BA.2–BA.4 were 1·70 and 1·95, corresponding to 76% and 74% cross-reactive responses, respectively. After accounting for waning, these equate to ∼36% (BA.1) and ∼35% (BA.2) of the initial boost at three months, and ∼18% and ∼17% at six months.

The posterior median estimate for the standard deviation of observed titres was 0·848 (95% CrI: 0·812–0·884), indicating substantial inter-individual variability and measurement noise (Table 1). For a true titre of five on the log_3_(titre/50) scale, there was ∼24% probability of observing a titre one log_3_ higher or lower, and only ∼2% for a deviation greater than two logs.

### Antibody kinetics is influenced by the infecting variant and participant response type

We next examined antibody kinetics stratified by infecting variant (pre-Omicron vs. Omicron) and response type (high vs. low responders) (Table 2). Pre-Omicron infections generated larger boosts overall, although the differences was smaller among high responders (Figs 2a,b). High responders consistently exhibited higher nAb titres than low responders, irrespective of variant (Figs 2c,d). Pre-Omicron exposures induced boosts of 2·434 (1·972-3·119) log_3_(nAb titre/50) units in high responders versus 0·859 (0·690-1·04) in low responders, while Omicron exposures induced boosts of 2·305 (1·896-2·824) in high responders but 0·413 (0·350-0·477) in low responders, showing that low responders infected with Omicron had the smallest nAb increase. Note that our model does not explicitly differentiate first-time infections from reinfections. The same boosting parameters were applied whether the infection was a first-time Omicron infection or a reinfection following pre-Omicron exposure. Since there were very few pre-Omicron infections in our cohort, these estimates likely mostly reflect first-time Omicron infections.

**Table 2.** Estimated values for the short-term boosting and cross reactivity parameters when stratified by infecting variant and individual response type, inferred using *serosolver*. The estimated value is on the log_3_(titre/50) scale.

| Parameter | Description | Units | Stratification | Estimate (posterior median; 95% CrI) – $\log_3(\text{titre}/50)$ |
| --- | --- | --- | --- | --- |
| $\mu_s$ | Short-term boosting | $\log_3(\text{titre}/50)$ units | Low responders, infected pre-Omicron | 0.859 (0.690-1.04) |
|  |  |  | High responders infected pre-Omicron | 2.434 (1.972-3.119) |
|  |  |  | Low responders, infected during Omicron | 0.413 (0.350-0.477) |
|  |  |  | High responders infected during Omicron | 2.305 (1.896-2.824) |
| $\sigma_s$ | Short-term cross reactivity | Proportion decrease in boost per unit of antigenic distance | Baseline – low responders, infected pre-Omicron | 0.155 (0.096-0.222) |
|  |  |  | High responders infected pre-Omicron | 0.012 (0.001-0.041) |
|  |  |  | Low responders, infected during Omicron | 0.005 (0.001-0.017) |
|  |  |  | High responders infected during Omicron | 0.002 (0.0001-0.023) |

CrI – credible interval.

**Fig 2.**
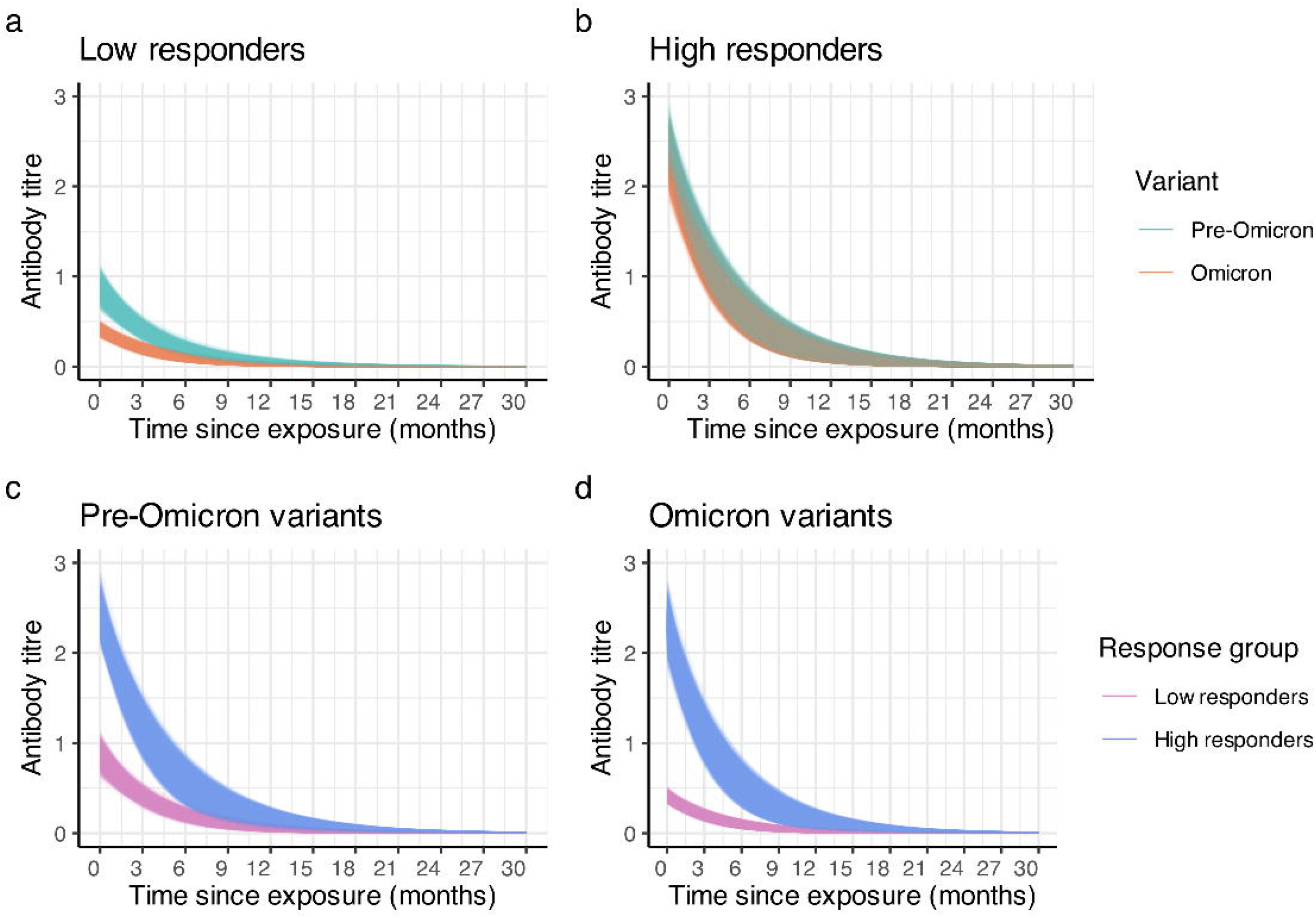
Estimated antibody kinetics stratified by infecting variant and response group. The x-axis shows time since exposure (months) and the y-axis log_3_(titre/50). Each line represents a single posterior draw. (A) Low responders: pre-Omicron (green) vs. Omicron (orange). (B) High responders: pre-Omicron vs. Omicron. (C) Pre-Omicron exposures: low (pink) vs. high responders (blue). (D) Omicron exposures: low vs. high responders.

Urban residence and age were both associated with a higher likelihood of being a high responder. Individuals living in urban areas were 1·65 times more likely to be high responders compared to those in rural areas (95% CI: 1·34–2·04; p=0·0014). Similarly, adults were 1·41 times more likely to be high responders than children (95% CI: 1·24–1·61; p=0·0021).

Cross-reactivity analyses showed broad short-term neutralisation across antigenic space in all subgroups (Fig 3). Omicron infections generally induced wider cross-neutralisation than pre-Omicron infections, while a decline in titre with increasing antigenic distance was most pronounced in pre-Omicron, low responders (Fig 3, Table 2). Overall, trajectories were similar across response types, but high responders maintained higher cross-neutralisation than low responders.

**Fig 3.**
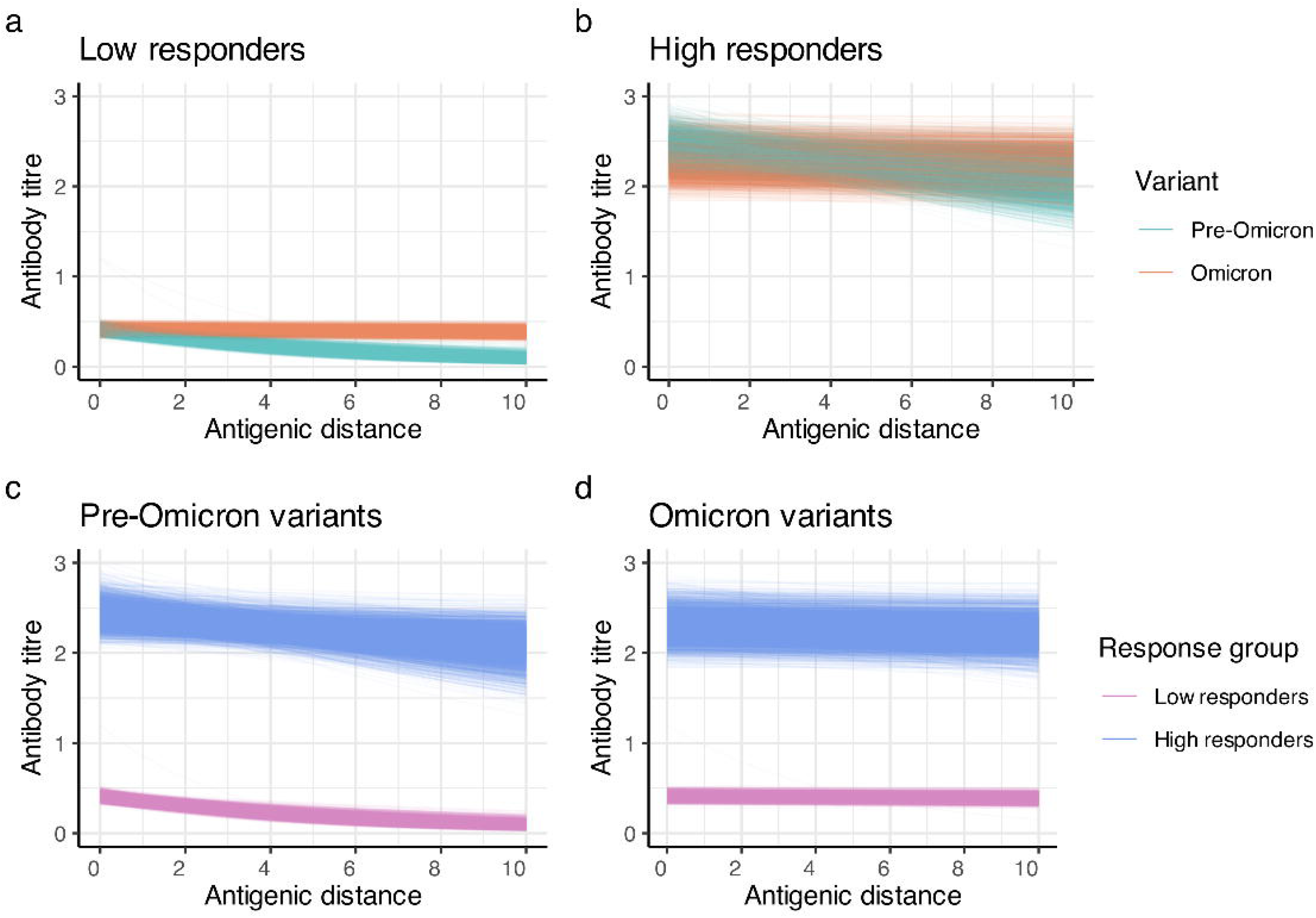
Antibody cross-reactivity stratified by infecting variant and response group. The x-axis shows antigenic distance between variants (1 unit on the antigenic map), and the y-axis log_3_(titre/50). Each line represents a single posterior draw. (A) Low responders: pre-Omicron (green) vs. Omicron (orange). (B) High responders: pre-Omicron vs. Omicron. (C) Pre-Omicron: low (pink) vs. high responders (blue). (D) Omicron: low vs. high responders.

### Inferred infection histories and individuals with repeat infections

*Serosolver* inferred individual infection histories from the longitudinal nAb data. Model fits from nine illustrative participants are shown, demonstrating the diversity of inferred trajectories (Fig 4). Some individuals showed single Omicron BA.1/BA.2 infections with modest antibody responses, others showed stronger post-Omicron responses, and some displayed evidence of earlier or repeat infections.

**Fig 4.**
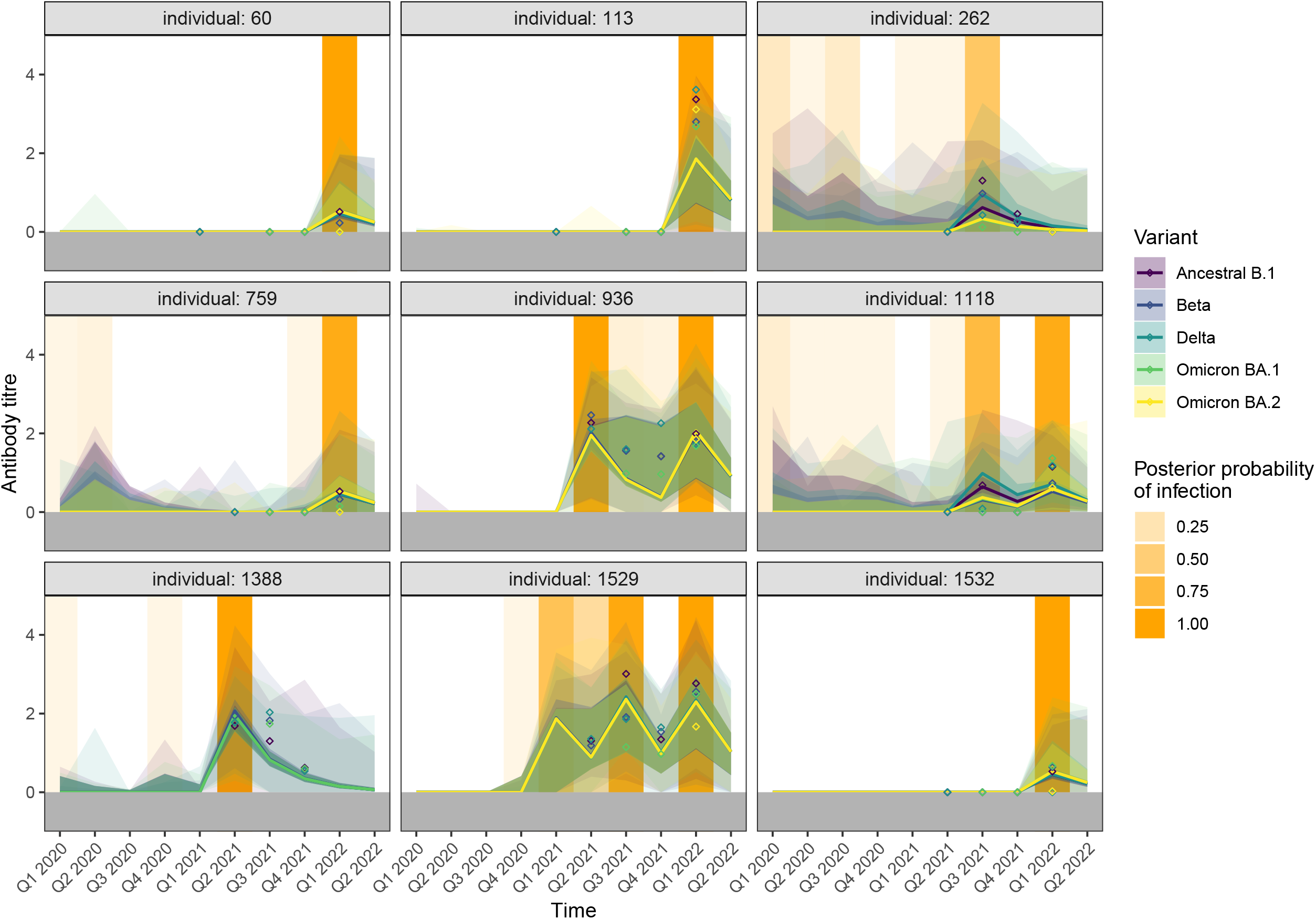
Modelled antibody trajectories for nine participants with inferred exposure events. Each panel represents an individual, with the x-axis showing time periods and the y-axis the log_3_(titre/50) antibody titre. Measured titres are indicated by diamonds, coloured by assay variant. Solid lines represent the modelled posterior mean trajectories, with shaded areas showing 95% prediction intervals. Orange bars indicate the posterior probability of infection at each time point, with darker shading representing higher probability. No BA.2 trajectory is shown for individual 943 due to the absence of a sample in the final survey (Q1 2022).

Repeat infections were inferred in 33 participants over the study period (Fig 5). Most experienced two infections (n=28), four had three, and one had four. Traditional serological analyses, based solely on seroconversion from seronegative to seropositive, would therefore have missed 39 reinfections, representing ∼9% of the 429 total infections identified by the model. The majority of reinfections (28/39; 71·8%) occurred during the Omicron BA.1/BA.2 wave in early 2022. The first infections in these individuals predominantly occurred in earlier waves, with 17 in Q1–Q2 2021, 13 in Q3 2021, and three in Q4 2021. The average interval between infections was approximately eight months. Reinfection risk was significantly higher among urban compared with rural participants (risk ratio 2·31, 95% CI 1·13–4·72, p=0·020) and among adults (≥15 years) compared with children (<15 years) (risk ratio 8·76, 95% CI 2·10–36·49, p<0·001). Reinfection risk was also significantly higher among the high responders, with 41% (12/29) of them experiencing reinfection compared to only 1·3% (21/1,646) of low responders, corresponding to a risk ratio of 32·4 (95% CI: 17·7–59·5, p<0·001).

**Fig 5.**
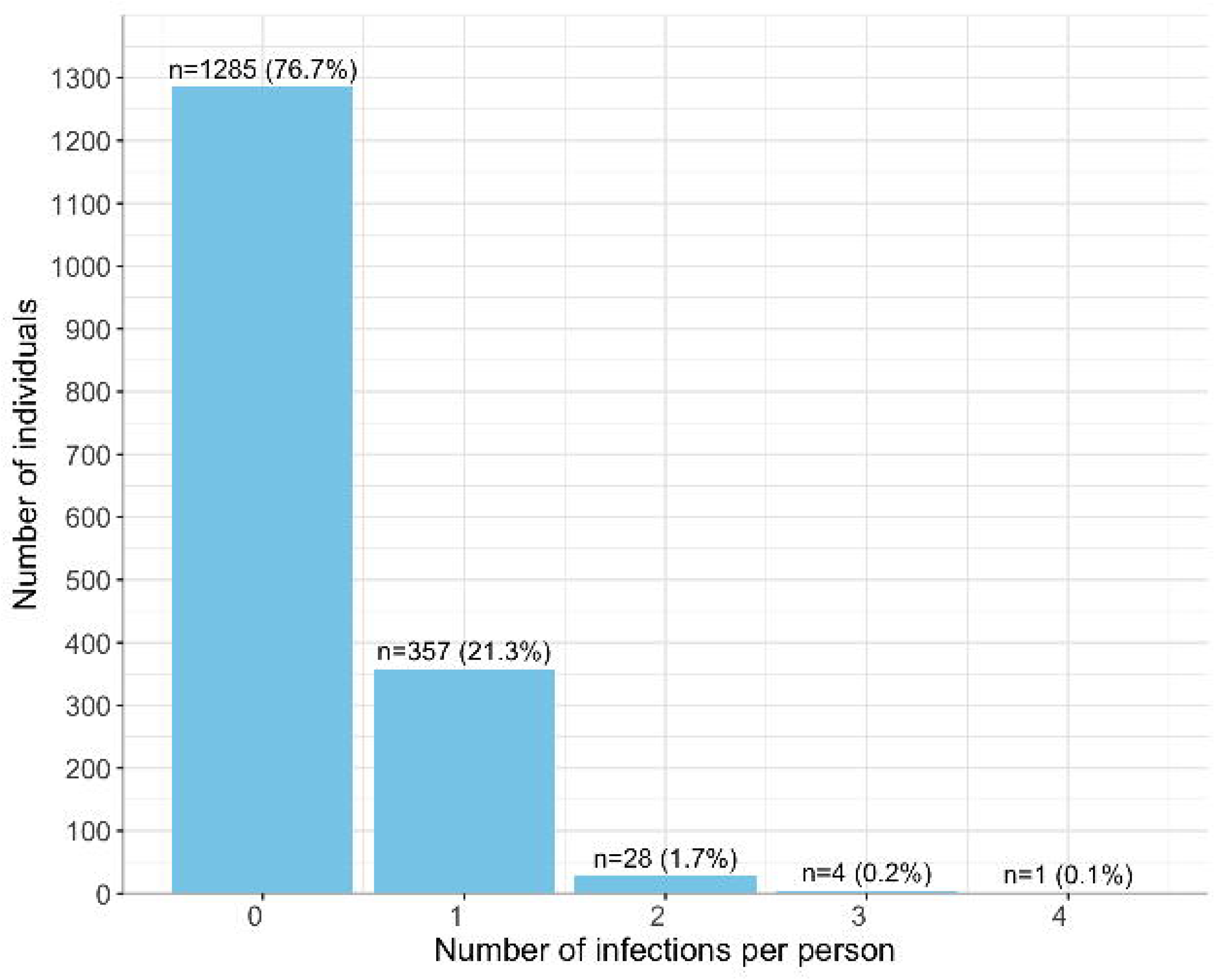
Distribution of multiple infection events among study participants. Bars show the number of individuals (y-axis) according to the posterior median number of infections per person (x-axis) across possible exposure periods from Q1 2020 to Q2 2022. Counts are shown with corresponding percentages. Note that this figure represents the overall distribution across the entire cohort; peak seroincidence at specific timepoints (reported in the text) can be higher because fewer participants remained in the study at later periods when most infections occurred.

### Seroincidence across the urban and rural regions of Malawi

Population-level infection rates (i.e. seroincidence) were derived from reconstructed individual infection histories, and were consistently higher in the urban region (Lilongwe) than in the rural region (Karonga) (Fig 6). Before Q1 2021, seroincidence rates (defined as the proportion of individuals with antibody increases detected by the model per quarter) were close to zero at both sites (posterior median estimates: 0·000–0·001; CrI 0·000–0·014). Modest increases were observed between Q2 and Q4 2021, coinciding with the Delta wave, although seroincidence remained below 10% in both sites. A sharp rise occurred in Q1 2022 during the Omicron BA.1/BA.2 wave, reaching a posterior median estimate of 0·41 (CrI: 0·39–0·43) in the urban region and 0·27 (CrI 0·25–0·28) in the rural region. Seroincidence in the rural area fell rapidly by Q2 2022 (posterior median estimate: 0·00, CrI 0·00–0·03). No samples were available for the urban area after Q1 2022.

**Fig 6.**
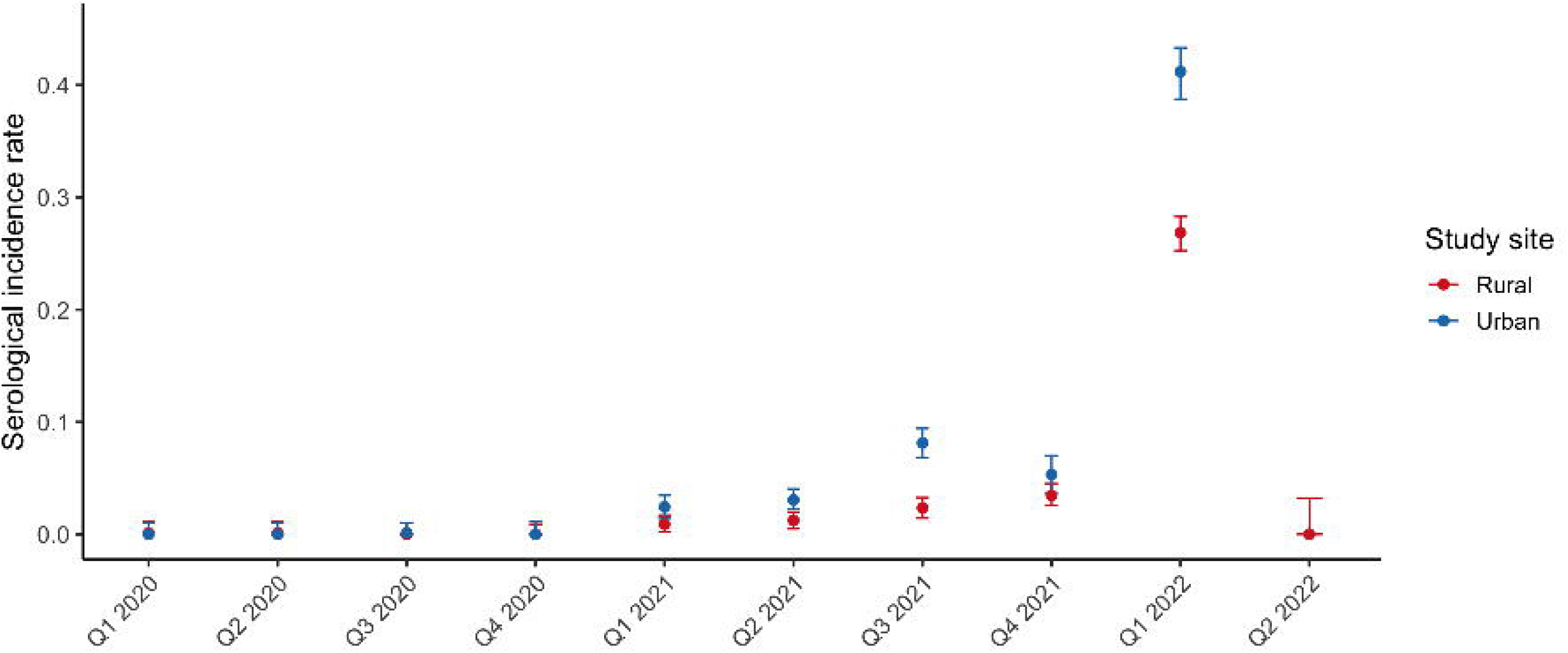
Estimated quarterly seroincidence of SARS-CoV-2 in the rural (Karonga, red) and urban (Lilongwe, blue) study sites from Q1 2020 to Q2 2022. The y-axis represents the seroincidence per capita per three months with a serological response detected by the model each quarter, and the x-axis indicates the time periods. Error bars show 95% credible intervals (CrI) around the posterior mean estimates. A pronounced rise in seroincidence was evident in Q1 2022, particularly in the urban population.

In contrast, traditional serology provided far less temporal resolution, capturing only the proportion of seroconverted individuals at each survey (S14 Fig), making it difficult to delineate epidemic waves or site-specific differences.

### Comparison of traditional serology with serosolver infection detection

Serosolver identified 429 infection events (95% CrI: 417–441), compared with 404 detected by traditional serological analyses. Most infections were identified by both approaches (88%, n=390/443). 39 additional reinfections (9% of all possible infections) were detected only by serosolver. Traditional serology identified 14 infections (3% of all possible infections) not inferred by *serosolver*; these individuals had very low posterior infection probabilities (<0.25), reflecting weak variant-specific nAb responses that the model attributed to measurement noise rather than true infections (S15 Fig).

### Influence of the antigenic map on the model

To assess the impact of antigenic map selection, we repeated the analysis using an alternative map. Estimated infection counts were nearly identical (posterior median 428, 95% CrI 417–440 vs. 429, 95% CrI 417–441), and seroincidence patterns remained unchanged (S16 Fig vs. Fig 6). Short-term boosting was slightly higher (0·989 vs. 0·859) and cross-reactivity decreased (0·386 vs. 0·115), reflecting lower neutralisation with higher antigenic distance (S3 Table vs. Table 1). Waning rates (0·707 vs. 0·734) and observation variability (0·821 vs. 0·848) were similar, indicating that the choice of antigenic map had minimal effect on infection detection or population-level inference.

## Discussion

Understanding population-level SARS-CoV-2 immunity remains essential in settings with limited routine surveillance [29,30]. Using longitudinal neutralisation data from 1,675 participants across urban and rural Malawi, we characterised antibody kinetics, infection histories, and estimated seroincidence over a 15-month period. Substantial heterogeneity in both exposure and immune responses were observed across individuals and study sites, reflecting the marked variability in SARS-CoV-2 transmission dynamics.

*Serosolver* inferred 429 infection events (95% CrI: 417–441), including 39 reinfections not detected by traditional serology, which detected 404 infections. These differences largely arose because traditional serology classified small antibody rises against a single variant as new infections, while *serosolver* integrates the totality of an individual’s antibody profile over time, accounting for cross-reactivity and measurement error. Estimated antibody boosting at the baseline was 0·859 log_3_(titre/50) (128·4 on the natural scale; CrI: 106·7– 156·9), aligns with estimates from studies in Hong Kong [31,32], although assay differences (PVNA vs. plaque reduction neutralisation titre (PRNT)90) limit direct comparison. Neutralising antibodies waned rapidly, with approximately half remaining at three months and only 5% at one year; modelled trajectories suggested near-complete loss by 18–24 months. Cross-neutralisation decreased with increasing antigenic distance (baseline decay rate of 0·155), with ancestral B.1 retaining ∼42% homologous activity against Omicron BA.1. These findings are consistent with Omicron’s extensive immune escape; however, it still remains somewhat susceptible to prior ancestral B.1 generated immunity. Using the measured antigenic distances, we estimated that Omicron BA.1/BA.2 infection offered limited short-term protection against BA.4, consistent with epidemiological evidence from the region [33].

Pre-Omicron variants elicited stronger but narrower neutralising responses, whereas Omicron infections produced broader yet weaker responses [27,34]. It is unclear whether this pattern reflects intrinsic properties of Omicron or cumulative prior exposures, as some individuals infected during the Omicron wave had experienced earlier infections and therefore immune imprinting. Broader responses may therefore reflect back-boosting of pre-existing memory B cells, which produce cross-reactive antibodies, alongside recruitment of new Omicron-specific antibody-secreting cells [35,36]. However, in our cohort very few people had been infected prior to the emergence of Omicron and therefore it is more likely that the difference is due to the intrinsic properties of Omicron.

High responders consistently mounted larger antibody boosts than low responders, reflecting substantial inter-individual heterogeneity potentially shaped by age, genetics, comorbidities, helminth co-infections [37,38], past exposure to coronaviruses, viral load, and lifestyle factors [39-41]. Categorising responses this way is meaningful, as these factors influence how individuals respond to infection. Distinguishing high and low responders can indicate susceptibility, and protection. At a population level, the balance of response types may affect viral spread, vaccine effectiveness, and herd immunity thresholds. Considering this variation in models and public health planning could improve outbreak predictions and help target interventions to those with weaker or stronger immune responses. Interestingly, it was found that being high responders was more frequent among adults and urban residents. Despite these significant relative differences, it is important to note that the overall proportion of high responders was low across all groups, meaning that the absolute differences in response rates remain small.

Reinfections were predominantly observed among individuals infected in early waves and among urban residents in Lilongwe, consistent with increased transmission in high-density settings. Similar patterns have been observed elsewhere, with dense urban populations experiencing a greater force of SARS-CoV-2 infection, compared to lower-density populations [42,43]. Moreover, the earlier emergence of the epidemic in urban settings (due to better connectivity of these regions with international travel), led to earlier immune waning, thereby rendering these individuals susceptible reinfection sooner. Reinfections were less frequently detected in children, which may indicate genuinely lower infection rates, or lower nAb titres that reduce the likelihood of reinfections being detected. Reinfections were more frequent among high responders, suggesting that repeated exposure strengthens nAb responses. Seroincidence peaked during the Omicron BA.1/2 wave in Q1 2022, mirroring findings from South Africa [44]. Pre-2021 infections were rare, consistent with limited early transmission in Malawi [9], although sparse data prevent definitive conclusions. Model outputs were robust to alternative antigenic maps, supporting the reliability of infection and seroincidence estimates despite uncertainty in antigenic positions.

The study has several strengths. Inclusion of both urban and rural cohorts enabled assessment of geographical heterogeneity in exposure. Longitudinal measurements of nAbs allowed reconstruction of infection timing, reinfections, and waning immunity, which is not possible with cross-sectional studies. Using nAb data strengthens the functional relevance of the inferred antibody kinetics, given their association with protection against severe disease. The identification of multiple infections improves estimates of transmission rates, population susceptibility, and the impact of variant immune escape. Importantly, these insights have practical public health implications: e.g. identifying groups at higher risk of reinfection, particularly urban adults and those with earlier infections, can support prioritisation of booster vaccinations, tailoring of public health messaging, and more effective deployment of limited surveillance resources.

Several limitations should be considered. First, the model assumes only one dominant variant per time period, homogeneous immune responses within subgroups, and symmetric cross-variant boosting. These simplifying assumptions may not fully reflect real-world transmission and immune dynamics. Stratification by high and low responders enabled separate parameter estimation, but thresholds were defined arbitrarily, and more flexible model-based approaches (latent classes for different responder types or random effects to capture individual-level variation in antibody responses) are not currently supported in *serosolver* [26]. Second, the antibody kinetics model structure and model describing the distribution of observed titres was chosen to balance immunological plausibility and model identifiability, but may not perfectly reflect true kinetics. Although modelling long- and short-term antibody responses as separate model parameters have been done previously [16,18], data scarcity precluded estimation of separate phases. Thus, the model was simplified to a single boosting and waning parameter. Furthermore, our longer term waning predictions are reliant on the chosen exponential waning model. Different function forms for antibody waning would lead to different predictions and therefore, long-term extrapolation should be interpreted with caution. Third, sampling occurred at three-month intervals across only four timepoints, limiting temporal resolution. Subgroup analyses by age, symptom severity, or healthcare attendance were underpowered, as evidenced by convergence issues in exploratory analysis. Vaccine-induced immunity could not be assessed as vaccinated individuals were excluded after their first vaccination visit, and individuals living with HIV were excluded from analysis, preventing examination of how immunosuppression affects antibody kinetics. The 90% neutralisation threshold may underestimate weaker responses, and assay variability may contribute additional noise.

Future work should incorporate longer follow-up to capture long-term antibody dynamics, explore determinants of immune variability, and link neutralisation titres to infection risk to refine correlates of protection. In line with recent longitudinal serosurveillance studies across the African region [20,21], our findings highlight the value of integrating serological and clinical data with mechanistic modelling to characterise population immunity in settings where routine surveillance is limited. Such approaches can provide more accurate, timely and actionable insights into evolving immunity and vulnerability.

In summary, we provided detailed characterisation of SARS-CoV-2 antibody kinetics and reinfection dynamics in urban and rural Malawian populations. Rapid waning of neutralising antibodies, partial cross-protection between variants, and higher urban exposure, underscores the need for continued vaccination and the value of high-resolution serological modelling in inform epidemic monitoring in resource-limited settings.

## Data Availability

All code used to generate the model and produce the figures is openly available at Github:
https://github.com/MJMcCormack/COVSERO_serosolver, together with accompanying documentation. The participant data are not publicly available because they contain information such as birth dates and location data, which could risk participant re-identification and would breach ethical and privacy requirements. Requests for access to individual-level, pseudo-anonymised data and supporting documents may be submitted to, referencing the paper title. Access will be subject to approval and completion of a data transfer agreement. Simulated versions of the data have been provided in the Github repository.

https://github.com/MJMcCormack/COVSERO_serosolver

## Acknowledgements

We extend our gratitude to all study participants, and the laboratory and field teams of the Malawi Epidemiology and Intervention Research Unit’s “COVSERO” study.

## Supplementary Information Captions

**S1 Supplementary Information. Supplementary Methods**. Additional details on laboratory procedures and *serosolver* model setup.

**S1 Table. Gene construct mutations for the different SARS-CoV-2 viruses, relative to the Wuhan-Hu-1 virus sequence (GenBank: MN908947).**

**S2 Table. *Serosolver* prior information.** This table provides an overview of all model parameters, including covariate coefficients (coef), the corresponding prior distributions, the parameters defining each prior (mean and standard deviation for Normal and Log-Normal distributions; alpha and beta for Beta distributions), and the permissible bounds for each parameter. Short-term boosting and cross-reactivity were stratified by response type to capture heterogeneity in individual-level antibody dynamics.

**S3 Table. Estimates of antibody kinetics parameters at baseline (low responders, infected pre-Omicron) inferred using the alternative antigenic map.** Values are expressed on the log_3_(titre/50) scale used in the model.

**S1 Fig. Map of Malawi indicating the study sites of Karonga Health and Demographic Surveillance Site (HDSS) (rural) and Area 25, Lilongwe (urban).**

**S2 Fig. Sample collection timeline for the study (February 2021–April 2022).** Samples were collected during four distinct survey rounds, each undertaken at separate time points, demonstrating the non-overlapping design of the data collection.

**S3 Fig. Schematic showing the multiple levels of the *serosolver* model.** (A) The model describes the probability of SARS-CoV-2 infection over time. (B) Infections are modelled as Bernoulli trials from the nfection probability model in (A), resulting in longitudinal neutralising antibody (nAb) titres. This example shows predicted antibody titres to variant X (blue) for a participant first infected with variant X and later with variant Y. (C) The model captures cross-reactive antibody response to all variants. Shown here are nAb titres to variant Y (pink) over time, compared to variant X (blue, dotted), with sampling points for two longitudinal measurements indicated. (D) The antigenic map determines cross-reactivity between variants, here depicting the Euclidean distance between variants X and Y. (E–F) nAb titres measured against variants X (blue) and Y (pink) at the first (E) and second (F) sample collection. Created with Biorender.com.

**S4 Fig. Observation model illustrating the distribution of predicted observations.** (A) Observation model for individuals with predicted titres above zero (i.e., individuals with at least one previous infection), showing exponential antibody waning and normally distributed measurements. (B) Observation model for individuals with predicted titres of zero (never infected), where true antibody levels are zero, but occasional false positives occur, represented by the false positivity rate *p*. Blue lines indicate true antibody trajectories over time (absent in B), while red lines show the corresponding observation distributions. LOD — limit of detection. Created with Biorender.com.

**S5 Fig. Alternative antigenic map used for sensitivity analysis.** Antigenic distances between SARS-CoV-2 variants are shown: ancestral B.1 (red), Beta (blue), Delta (green), Omicron BA.1 (purple), and Omicron BA.2 (orange). Map coordinates were derived from Mykytyn et al. 2023 [13].

**S6 Fig. Beta(1/3, 1/3) prior distribution.** The x-axis shows the probability of infection, and the y-axis shows the corresponding density. Created with Biorender.com.

**S7 Fig. SARS-CoV-2 neutralising antibody (nAb) titres by variant across the study sites and surveys.** (A) Rural region (Karonga) (B) Urban region (Lilongwe). Antibody titres against each of the SARS-CoV-2 variants were measured using an HIV(SARS-CoV-2) pseudotyped virus neutralisation assay and transformed using log_3_(titre/50) to match the dilution range. Omicron BA.1 and BA.2 were not tested against at Survey 1 as it had they not yet emerged, while Omicron BA.2 was only tested against at Survey 4. Boxplots show median and IQR of 90% titres. The whiskers stretch to the farthest data points that lie within 1.5 × IQR of Q1 and Q3, while any points beyond this range are displayed as outliers.

**S8 Fig. Individual neutralising antibody profiles for three example participants across the study period.** Antibody titres against each of the SARS-CoV-2 variants were measured using an HIV(SARS-CoV-2) pseudotyped virus neutralisation assay and transformed using log_3_(titre/50) to match the dilution range.

**S9 Fig. Serosolver trace plots demonstrating good convergence for the parameters in the final version of the model.** Three chains are shown: Chain 1 (red), Chain 2 (green), and Chain 3 (blue). The x-axis represents iteration number, and the y-axis shows parameter values.

**S10 Fig. Serosolver density plots demonstrating good convergence. Density plots for three chains (Chain 1: red, Chain 2: green, Chain 3: blue) show the posterior distributions of model parameters.** The x-axis represents parameter values, and the y-axis shows density. Strong convergence is indicated by overlapping density curves across chains, confirming that each chain sampled consistently from the same posterior distribution and that estimates are stable and independent of starting values.

**S11 Fig. Posterior predictive checks showing observed vs. predicted antibody measurements.** (a) Undetectable titres. (b) Non-zero titres. Histograms comparing observed antibody titres (orange) with predicted titres from the model (light blue) based on randomly selected posterior draws. The x-axis shows antibody titres (log_3_(titre/50) scale) and the y-axis shows the number of measurements at each level. Each panel represents a different posterior sample, and the close match between observed and predicted distributions indicates that the model captures the main features of the data well. Note that zero and non-zero titres are shown separately due to the vastly different y-axis scales.

**S12 Fig. Observed vs. predicted antibody titres.** Each point shows the observed antibody titre (black) and the model-predicted value (blue) with 95% prediction intervals. The x-axis represents individual measurement IDs sorted by observed titre, and the y-axis shows titres on a log_3_(titre/50) scale. The model accurately predicts most negative measurements, though a few observed negatives correspond to low predicted positives, likely reflecting weak true antibody responses that fall below the detection limit.

**S13 Fig. Modelled trajectories of neutralising antibody (nAb) titres following infection.** Antibody titres (log_3_(titre/50)) are plotted against time since exposure (months). The infecting variant (variant X, blue) and a cross-reactive variant (variant Y, pink) are shown. Variant Y lies 3.30 antigenic units from variant X, resulting in approximately a 40% lower boost relative to the homologous response.

**S14 Fig. Proportion of participants experiencing first-time seroconversion across four sequential surveys in the rural (Karonga, orange) and urban (Lilongwe, green) sites.** Bars indicate the fraction of participants who seroconverted in each survey, with 95% binomial confidence intervals shown. Surveys were conducted as follows: Survey 1 (24 Feb–8 Jun 2021), Survey 2 (28 Jun–13 Sep 2021), Survey 3 (4 Oct–10 Dec 2021), and Survey 4 (27 Jan– 22 Apr 2022). A marked increase in seroconversion is evident in Survey 4.

**S15 Fig. Model fits for 14 participants with infections detected only by traditional serology.** Each panel represents an individual. The x-axis shows time periods, and the y-axis shows antibody titre on the log_3_(titre/50) scale. Diamonds indicate measured titres, coloured by assay variant. Solid lines show posterior mean model trajectories, with shaded areas representing 95% prediction intervals. Orange bars indicate the posterior probability of infection, with darker shading denoting higher probability. Participant 113 is included for comparison to a strong serosolver-positive individual. Note that for individuals 865 and 1406, the measured titres are only just above zero (on the log_3_(titre/50) scale) for one of the variants. This would still count as a positive with traditional serology.

**S16 Fig. Estimated quarterly seroincidence of SARS-CoV-2 in the rural (Karonga, red) and urban (Lilongwe, blue) study sites from Q1 2020 to Q2 2022 – using the alternative antigenic map.** The y-axis represents the seroincidence per capita per three months, and the x-axis indicates the time periods. Error bars show 95% credible intervals (CrI) around the posterior mean estimates.

## Notes

### Competing Interest Statement

JAH has received consulting fees from Gerson Lehrman Group (GLG). All other authors declare no competing interests.

### Funding Statement

This research was funded by the Wellcome Trust (grants 217073/Z/19/Z awarded to AC, 221989/Z/20/Z awarded to AC and AH, and a Wellcome Trust Early Career Award 225001/Z/22/Z awarded to JAH), and by the Medical Research Council (grant MC_UU_00034/6 awarded to AH). The funders had no role in study design, data collection and analysis, decision to publish, or preparation of the manuscript.

### Author Declarations

Ethical approval for this study was obtained from the Malawi College of Medicine Research Ethics Committee (P11/20/3177, approved on 11 December 2020) and the University of Glasgow College of Medicine, Veterinary and Life Sciences Research Ethics Committee (200200056, approved on 8 February 2021).

