## Supplementary material for "SARS-CoV-2 neutralising antibody profiles reveal variant specific antibody dynamics and regional differences in infection histories in Malawi": S1 Supplementary Information

### Laboratory protocols

#### Cells

All cell lines were cultured at 37 °C in a humidified atmosphere containing 5% CO₂. Human embryonic kidney (HEK) 293-angiotensin converting enzyme 2 (ACE2) cells, generated via stable transduction of HEK293 cells with the pSCRPSY-hACE2 construct encoding the human ACE2 receptor, were kindly provided by Matt Turnbull and Suzannah Rihn (MRC–University of Glasgow Centre for Virus Research). These cells were cultured in Dulbecco’s Modified Eagle’s Medium (DMEM; Gibco) supplemented with 10% foetal bovine serum (FBS; Gibco), 2 mM L-glutamine (Gibco), 100 IU/ml penicillin, and 100 µg/ml streptomycin (Gibco), hereafter referred to as complete DMEM. To maintain selection for ACE2 expression, 2 µg/ml puromycin (Merck Life Science UK Limited) was added to the culture medium. HEK293T cells [1], obtained from the American Type Culture Collection (ATCC) via University College London, were also maintained in complete DMEM, with the addition of 400 µg/ml G418 to preserve stable gene expression.

#### Generation of HIV(SARS-CoV-2) pseudotypes

HEK293T cells were transfected with plasmids encoding the SARS-CoV-2 spike protein corresponding to various variants—ancestral B.1, Alpha, Beta, Delta, Omicron BA.1, and Omicron BA.2—at a concentration of 0**·**15 µg/ml. Co-transfections included the HIV gag-pol plasmid p8.91 (0**·**1 µg/ml) and the firefly luciferase reporter plasmid pCSFLW2 (0**·**1 µg/ml), using polyethylenimine (PEI, 1 µl/ml; Polysciences, Warrington, USA) as the transfection reagent. Supernatants containing HIV(SARS-CoV-2) pseudotyped particles were collected 48 hours after transfection, filtered through a 0**·**45 µm membrane to remove cellular debris, aliquoted, and stored at −80 °C until further use. All spike gene constructs were codon-optimised and synthesised by GenScript Biotech. They included the ancestral B.1 (D614G), Beta (B.1.351), Delta (B.1.617.2), Omicron BA.1 (B.1.1.529), and Omicron BA.2 sequences (S1 Table). These constructs served as the basis for generating the variant-specific pseudoviruses used in neutralisation assays.

#### HIV(SARS-CoV-2) PVNA single dilution screening

Serum samples and controls were initially diluted 1 in 25 in complete DMEM and added in duplicate (25 µl per well) to white 96-well plates, followed by incubation with an equal volume of the relevant pseudovirus variant at 37 °C for one hour. HEK293-ACE2 target cells (4×10⁵ cells/ml, 50 µl per well) were then added, and plates were incubated at 37 °C for 48–72 hours. Luciferase activity was quantified using Steadylite Plus chemiluminescence substrate (75 µl, 1 in 3 dilution; Revvity, Beaconsfield, UK) and measured on an EnSight multimode plate reader (Revvity). Samples demonstrating >90% neutralisation were considered positive. Assay controls included a no-serum control (complete DMEM), a positive control (pooled SARS-CoV-2-positive sera collected March–May 2020, NHS Greater Glasgow and Clyde Biorepository, application 550), and a negative control (pre-pandemic serum from a single donor, Scottish National Blood Transfusion Service, protocol NATF 765 10).

#### HIV(SARS-CoV-2) PVNA titration

Serum titrations were performed using 3-fold serial dilutions from 1 in 25 to 1 in 18,225 in complete DMEM. Diluted sera (25 µl, in triplicate) were incubated with the pseudovirus (25 µl per well) for one hour at 37 °C, followed by addition of HEK293-ACE2 cells (4×10⁵ cells/ml, 50 µl per well). Plates were incubated for 38–72 hours before measuring luciferase activity as above. Neutralising titres were defined as the serum dilution achieving 90% reduction in infectivity. The viruses tested against were specified by study survey, as follows:

- Survey 1 – ancestral B.1, Beta, Delta
- Survey 2 – ancestral B.1, Beta, Delta, Omicron BA.1
- Survey 3 – ancestral B.1, Beta, Delta, Omicron BA.1
- Survey 4 – ancestral B.1, Beta, Delta, Omicron BA.1, Omicron BA.2

### *Serosolver* model details

#### Observation model

In the Hay et al. (2020) serosolver framework [2], log antibody titres measured at time $t$ are modelled as draws from a discretised, censored normal distribution centred on the latent “true” titre $A_{i,j,t}$ with variance $\varepsilon^{2}$. Here, $A_{i,j,t}$ represents the expected titre for individual $i$ following a potential infection at time $j$ and measured at time $t$. The model explicitly handles the assay’s lower and upper limits of detection (LODs). The likelihood of observing titre $Y_{i,j,t}\in\{0,\ldots,q_{\max}\}$ is:

$$P\left( Y_{i,j,t}|Z_{i},\theta\right)=f\left( Y_{i,j,t}|A_{i,j,t} \right)=\left\{ \begin{aligned} \int_{Y_{i,j,t}}^{Y_{i,j,t}+1} g(s)ds if Y_{i,j,t}\in\left\{ 1,q_{max}-1 \right\} \\ \int_{-\infty}^{1} g(s)ds if Y_{i,j,t}=0 \\ \int_{q_{max}}^{\infty} g(s)ds if Y_{i,j,t}=q_{max} \end{aligned} \right.$$

where $g(s)=\frac{1}{\sqrt{2\pi\varepsilon}}e^{-\frac{{(q-y_{i,j})}^{2}}{2\varepsilon}}$ denotes the probability density function of the normal distribution with mean $A_{i,j,t}$ and variance ε. Hence, $q_{max}$ denotes the maximum detectable titre (i.e. the upper limit of the assay).

Observed titres are thus treated as interval-censored realisations of a continuous distribution: $Y=0$ corresponds to titres below the detection limit and $Y=q_{\max}$ to titres beyond the assay’s range.

Because our dataset comprised continuous measurements with many observations at the lower limit, we modified the original observation model to handle continuous data and an excess of negative titres.

For individuals with non-zero predicted titres (i.e., previously infected at least once), the censored-normal formulation was retained but simplified by removing interval integration (S4a Fig):

$$P\left( Y_{i,j,t}|Z_{i},\theta\right)=f\left( Y_{i,j,t}|A_{i,j,t} \right)=\left\{ \begin{aligned} \int_{Y_{i,j,t}}^{Y_{i,j,t}+1} g(s)ds if Y_{i,j,t}\in\left\{ 1,q_{max}-1 \right\} \\ g(s)ds if Y_{i,j,t}=0 \\ \int_{q_{max}}^{\infty} g(s)ds if Y_{i,j,t}=q_{max} \end{aligned} \right.$$

This allows the model to capture both measurable titres and undetectable values arising from assay noise or low antibody concentrations.

For participants with no prior infection (true titre = 0), we introduced a false-positivity component absent from the original framework:

$$P\left( Y_{i,j,t}|never infected \right)=\left\{ \begin{aligned} 1-p if Y_{i,j,t} =0 \\ \frac{p}{q_{max}} if Y_{i,j,t}>0 \end{aligned} \right.$$

where $p$ is the false-positive rate. Based on the two-stage neutralisation assay—95 % specificity in the screening step and 97 % in the titration step—the combined false-positive probability was assumed to be:

$$p_{\text{overall}}=(1-0.95)\times(1-0.97)=0.0015.$$

Hence, false positives among truly seronegative individuals were rare, consistent with the high assay specificity, and assumed to be uniformly distributed within the detectable range (S4b Fig).

#### Stratification of parameters

We updated the *serosolver* model to allow parameters to be stratified by one or more categorical variables. Each stratum’s parameter values were estimated relative to a baseline group using a regression framework, for example:

$$\begin{matrix} \text{Baseline group 0:} & \mu_{s}^{(0)}=\mu_{s} \\ \text{Stratum 1:} & \mu_{s}^{(1)}=e^{{log(\mu}_{s})+\beta_{1}} \\ \text{Stratum 2:} & \mu_{s}^{(2)}=e^{{log(\mu}_{s})+\beta_{2}} \end{matrix}$$

and similarly for additional strata. The link function depends on the parameter’s bounds:

- Unbounded parameters: identity link, $\alpha_{j}=\alpha+\beta_{j}$
- Non-negative parameters (≥0): log link, $\alpha_{j}=e^{log(\alpha)+\beta_{j}}$
- Bounded parameters (0–1): logistic link, $\alpha_{j}=\text{logistic}\left( \text{logit}\left( \alpha\right)+\beta_{j} \right)$

#### Antigenic map sensitivity analysis

To assess the influence of the underlying antigenic map on model outputs, we re-ran the analysis using an alternative published map from Mykytyn et al. 2023 (S5 Fig) [3]. This map, generated from experimentally infected hamsters and measured via plaque reduction neutralisation assay (PRNT) against a panel of SARS-CoV-2 variants, differs in antigen positioning and relative distances compared with our primary map. By comparing inferred antibody kinetics parameters and seroincidence rates between maps, we evaluated the sensitivity of model outputs to these differences. This exercise provided a more robust validation of our findings and quantified the uncertainty arising from discrepancies among published antigenic cartography datasets.

#### Infection history prior details

We specified priors on infection histories using serosolver’s Beta–Bernoulli framework. Each individual’s infection history is a vector of 0s and 1s, with each index representing a three-month window (1 = infection, 0 = no infection). Infections are constrained so that individuals cannot be infected before birth or after their last serum sample.

In the Beta–Bernoulli model, the infection probability for each time window is drawn from a Beta distribution. We used a flexible Beta(1/3, 1/3) prior (S6 Fig), which is U-shaped, placing density near 0 and 1. This allows for sparse or clustered infections and accommodates large variation in infection probabilities across windows, similar in effect to Jeffreys’ prior. The Bernoulli component links individual infection probability to the population-level probability, making the framework equivalent to a Beta–Binomial distribution. This captures dependencies between individuals’ infection states while retaining flexibility in attack rates over time. Furthermore, because the Beta-Bernoulli prior effectively pools infection probabilities from all individuals, we modified *serosolver* to assume that individuals in different locations (urban vs. rural) followed separate Beta-Bernoulli distributions. This allowed to model to better detect different force of infection estimates between locations.

We also explored alternative priors to assess sensitivity. Using a Beta(1,10) prior produced slightly lower attack rates during the BA.1 period and higher rates elsewhere, but total infections per individual were largely unchanged. This occurs because Beta(1/3, 1/3) allows for large fluctuations in infection probability across windows, whereas Beta(1,10) smooths inferred rates over time without substantially affecting cumulative infections.
