## Supplementary material for "SARS-CoV-2 neutralising antibody profiles reveal variant specific antibody dynamics and regional differences in infection histories in Malawi": S1 Table

### **S1 Table. Gene construct mutations for the different SARS-CoV-2 viruses, relative to the Wuhan-Hu-1 virus sequence (GenBank: MN908947).**

| SARS-CoV-2 virus | Mutations relative to Wuhan-Hu-1 sequence |
| --- | --- |
| Ancestral (B.1) | D614G |
| Beta (B.1.351) | D80A, D215G, L241del, L242del, A243del, K417N, E484K, N501Y, D614G, A701V |
| Delta (B.1.617.2) | T19R, G142D, Δ156-157, R158G, L452R, T478K, D614G, P681R, D950N |
| Omicron BA.1 (B.1.1.529) | A67V, Δ69–70, T95I, G142D/Δ143–145, Δ211/L212I, ins214EPE, G339D, S371L, S373P, S375F, K417N, N440K, G446S, S477N, T478K, E484A, Q493R, G496S, Q498R, N501Y, Y505H, T547K, D614G, H655Y, N679K, P681H, N764K, D796Y, N856K, Q954H, N969K, L981F |
| Omicron BA.2 | T19I, △24/26, G142D, V213G, G339D,S371F, S373P, S375F, T376A,  D405N, R408S, K417N, N440K, S477N, T478K, E484A, Q493R, Q498R,  N501Y, Y505H, D614G, H655Y, N679K, P681H, N764K, D796Y, Q954H, N969K |
