## Supplementary material for "SARS-CoV-2 neutralising antibody profiles reveal variant specific antibody dynamics and regional differences in infection histories in Malawi": S2 Table

**S2 Table. *Serosolver* prior information.** This table provides an overview of all model parameters, including covariate coefficients (coef), the corresponding prior distributions, the parameters defining each prior (mean and standard deviation for Normal and Log-Normal distributions; alpha and beta for Beta distributions), and the permissible bounds for each parameter. Short-term boosting and cross-reactivity were stratified by response type to capture heterogeneity in individual-level antibody dynamics.

| Parameter | Description | Distribution | Prior values | Bounds | Stratification |
| --- | --- | --- | --- | --- | --- |
| coef | Model coefficients | Normal | Mean = 0, SD = 2 | NA | NA |
| µ_s_ | Short-term boosting | Log-normal | Log-mean = log(2),  log-SD = 1 | [0,8] | Response type, pre-Omicron vs Omicron exposures |
| σ_s_ | Short-term cross reactivity | Beta | Alpha = 2, Beta = 8 | [0,1] | Response type, pre-Omicron vs Omicron exposures |
| ω | Waning rate parameter for the short-term response (per three months) | Beta | Alpha = 1, Beta = 1 | [0,1] | Pre-Omicron vs Omicron exposures |
| ε | Standard deviation of observations | Normal | Mean = 0, SD = 2 | [0,25] | NA |

SD – standard deviation; NA – not applicable.
