## Supplementary material for "SARS-CoV-2 neutralising antibody profiles reveal variant specific antibody dynamics and regional differences in infection histories in Malawi": S3 Table

**S3 Table. Estimates of antibody kinetics parameters at baseline (low responders, infected pre-Omicron) inferred using the alternative antigenic map.** Values are expressed on the log₃(titre/50) scale used in the model.

| Parameter | Description | Estimate (posterior median; 95% CrI) – log_3_(titre/50) |
| --- | --- | --- |
| µ_s_ | Short-term boosting | 0**·**989 (0**·**842-1**·**16) |
| σ_s_ | Short-term cross reactivity | 0**·**386 (0**·**291-0**·**492) |
| ω | Waning rate parameter for the short-term response (per three months) | 0**·**707 (0**·**578-0**·**863) |
| ε | Standard deviation of observations | 0**·**821 (0**·**787-0**·**856) |

CrI – credible interval.
