## Supplementary figures and images for "SARS-CoV-2 neutralising antibody profiles reveal variant specific antibody dynamics and regional differences in infection histories in Malawi"

### S1 Fig

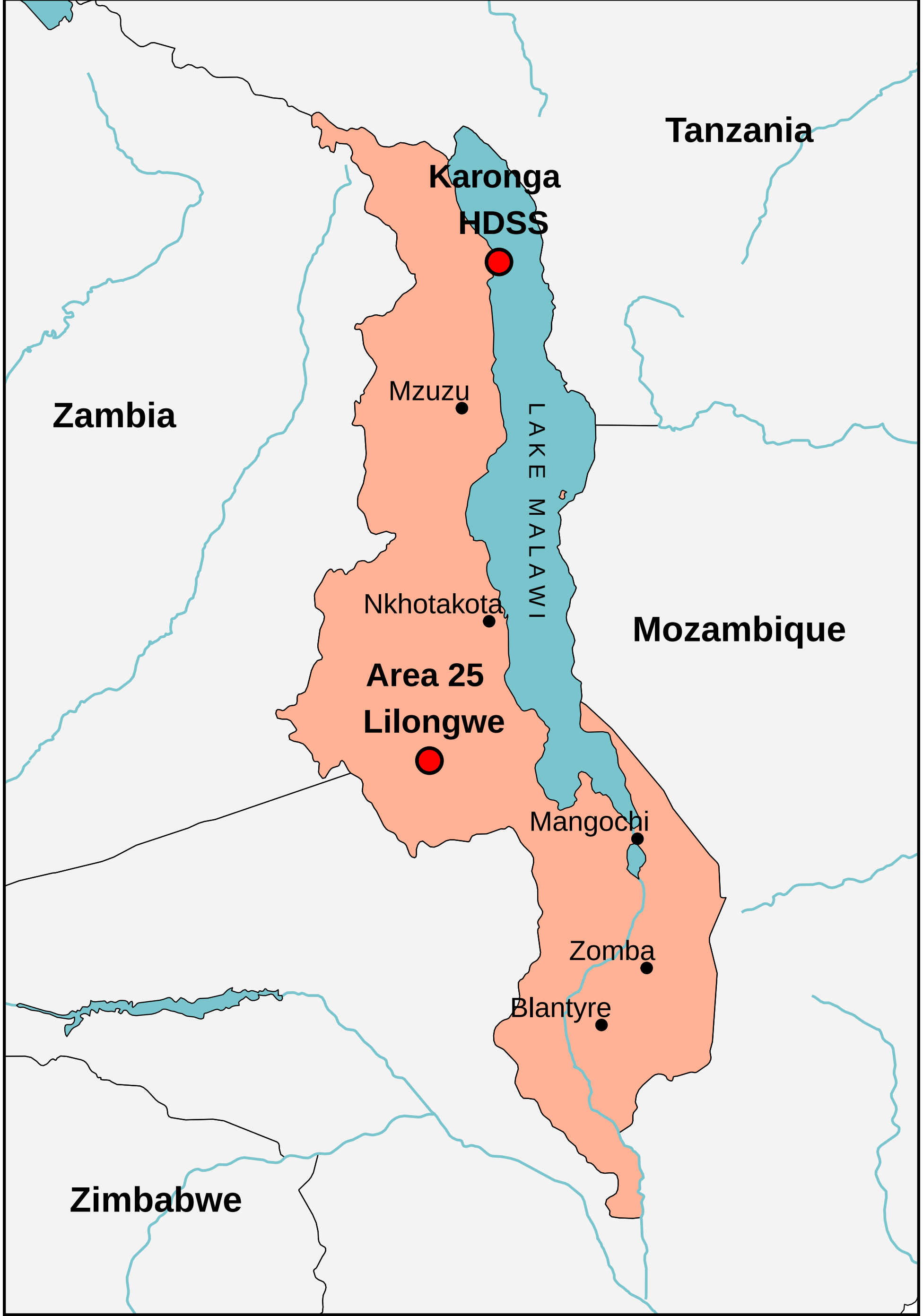

### S2 Fig

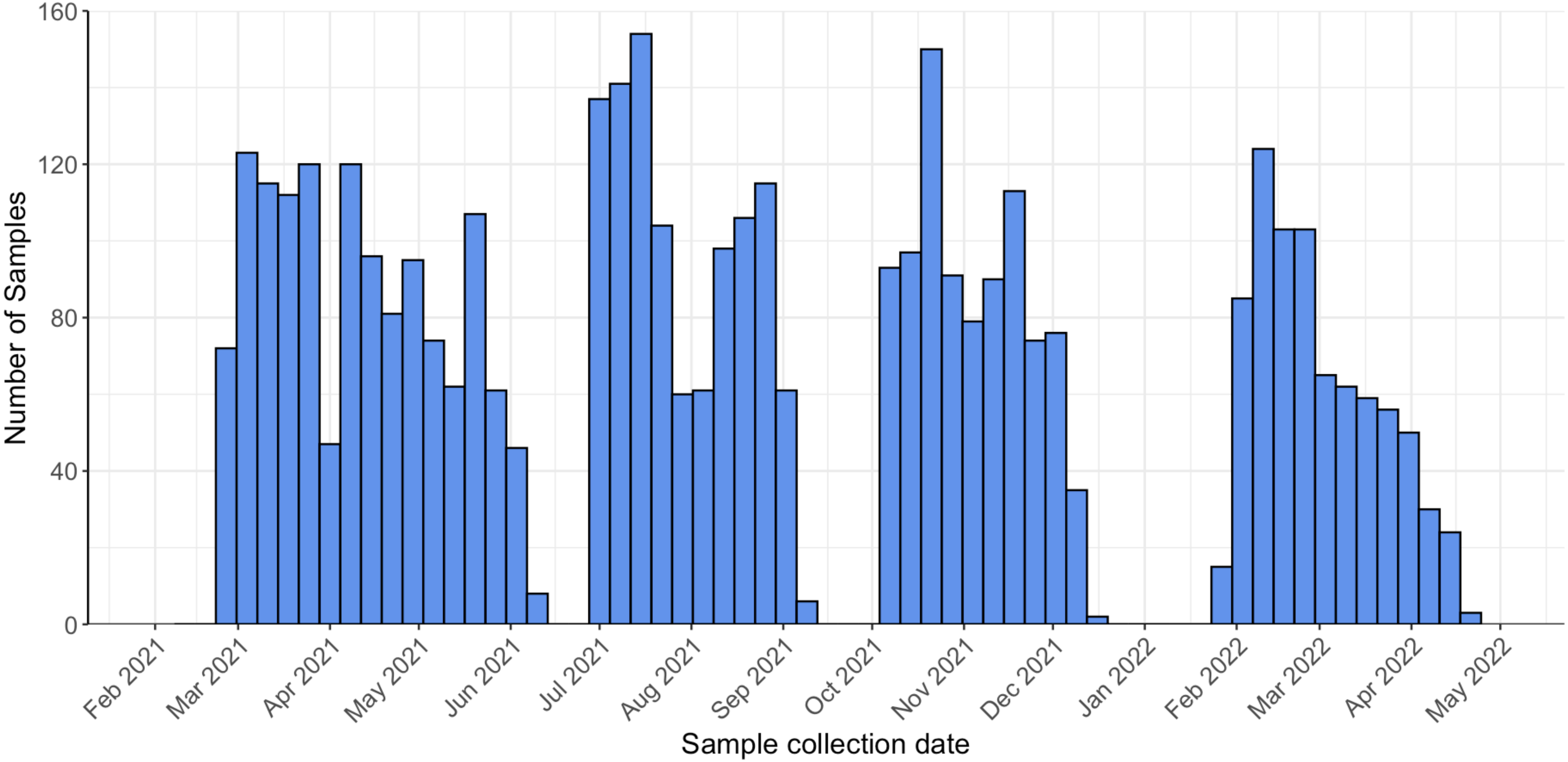

### S3 Fig

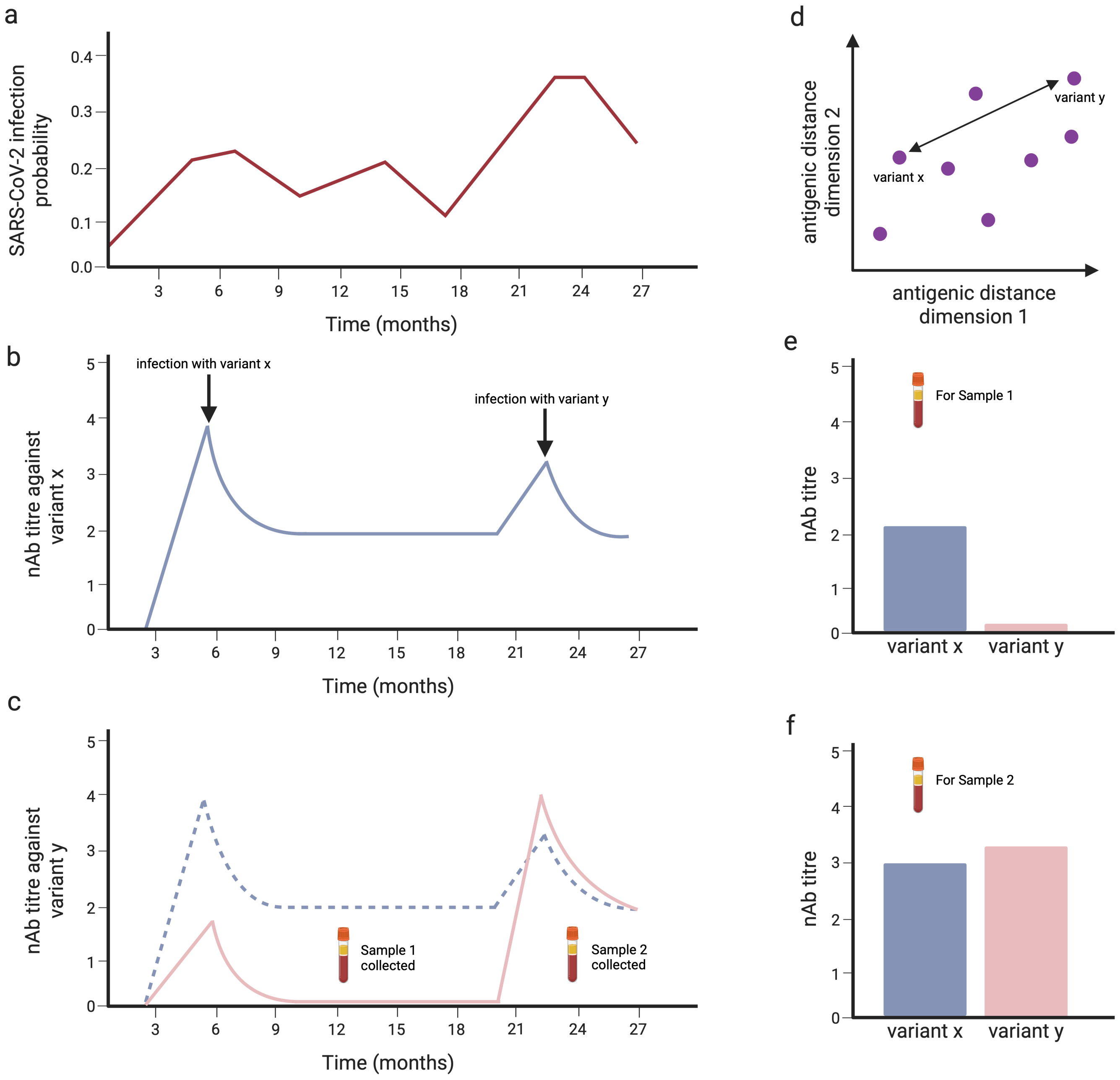

### S4 Fig

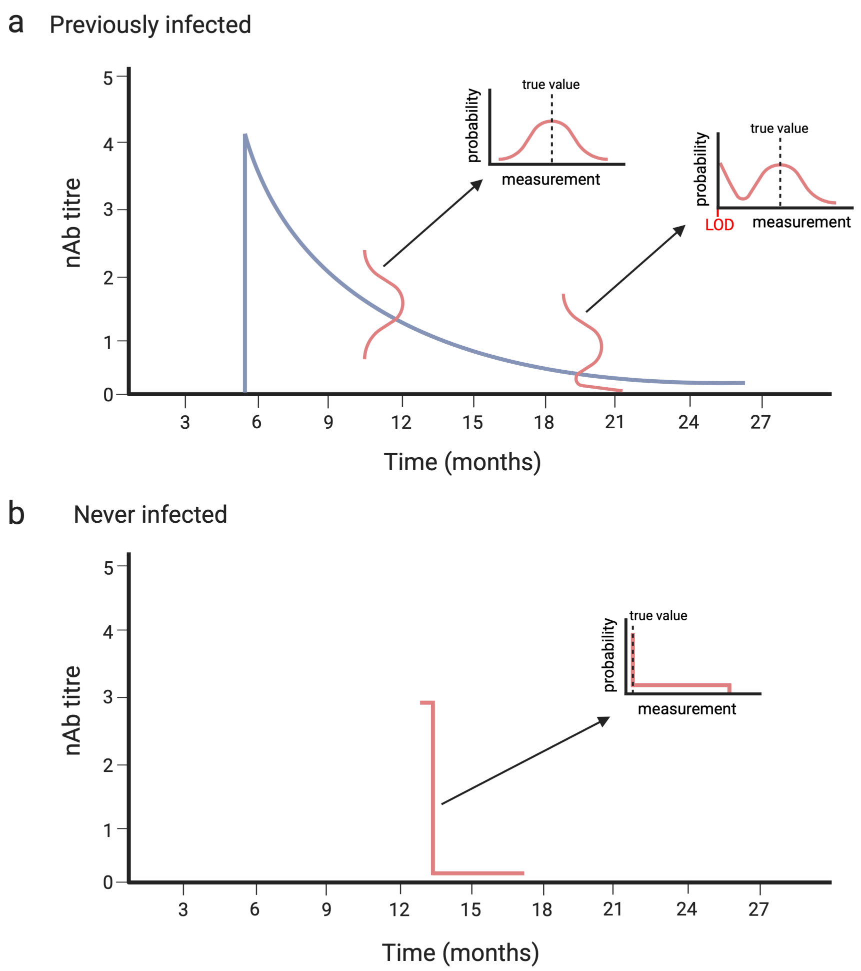

### S5 Fig

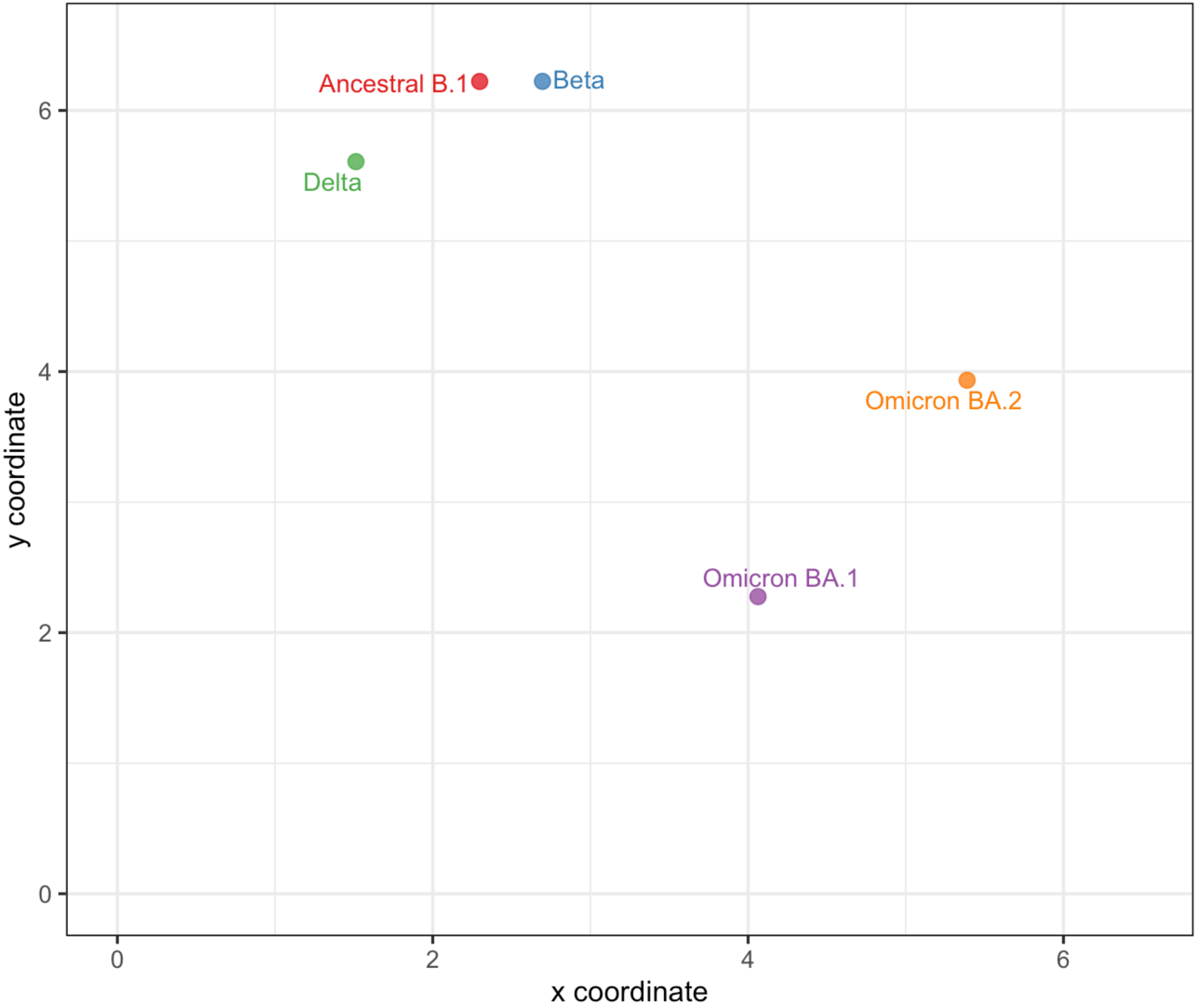

### S6 Fig

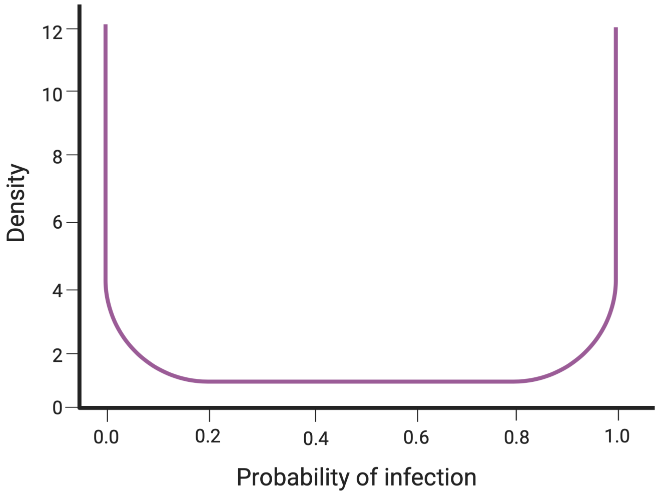

### S7 Fig

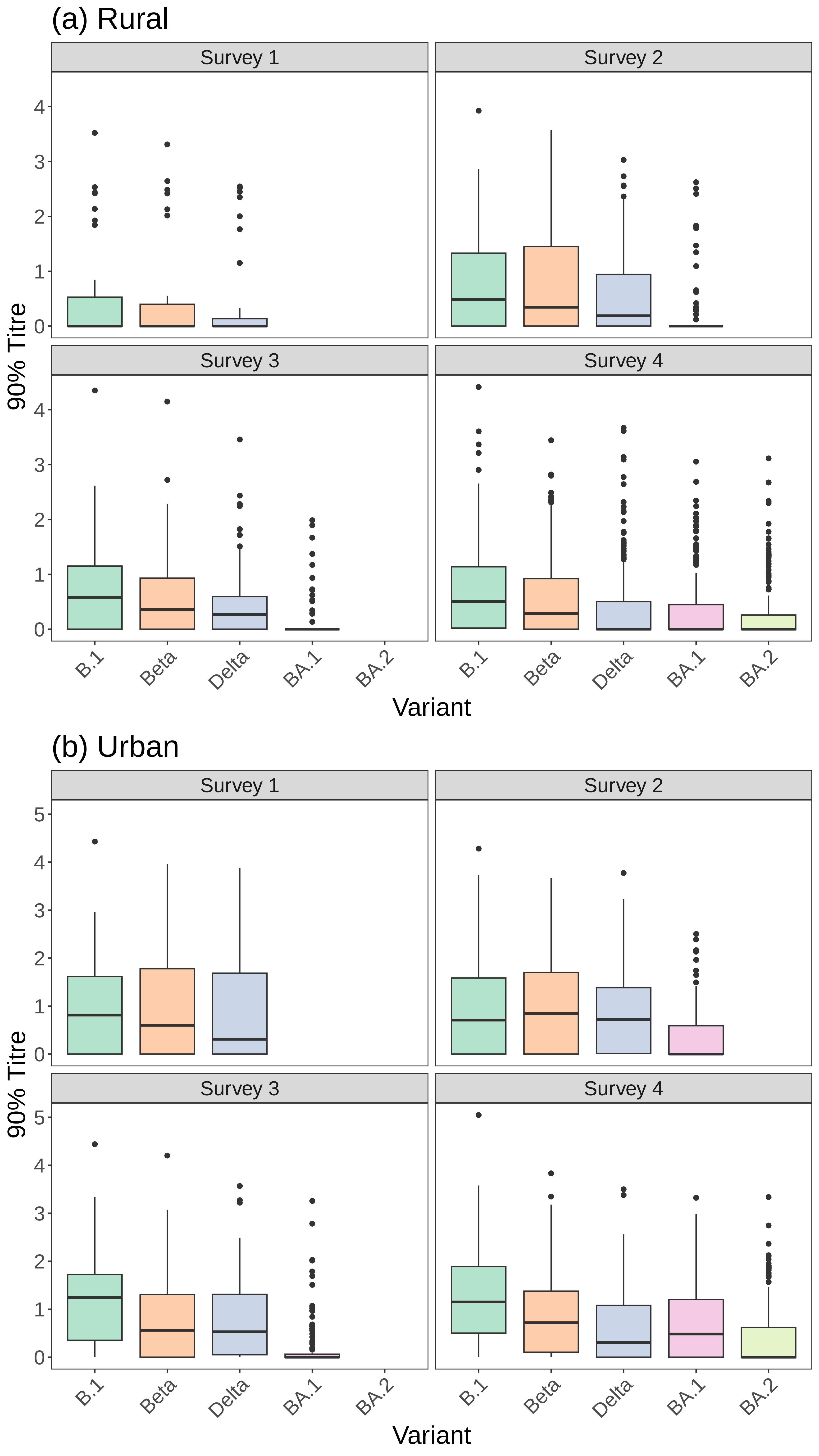

### S8 Fig

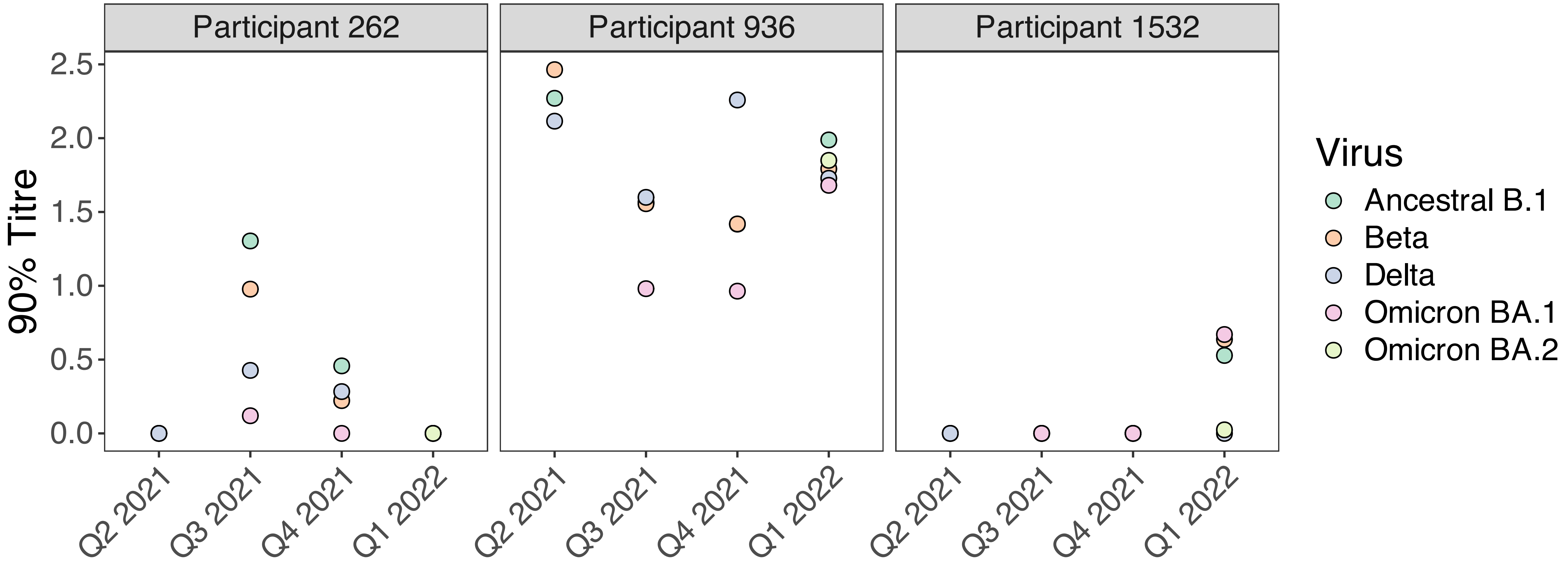

### S9 Fig

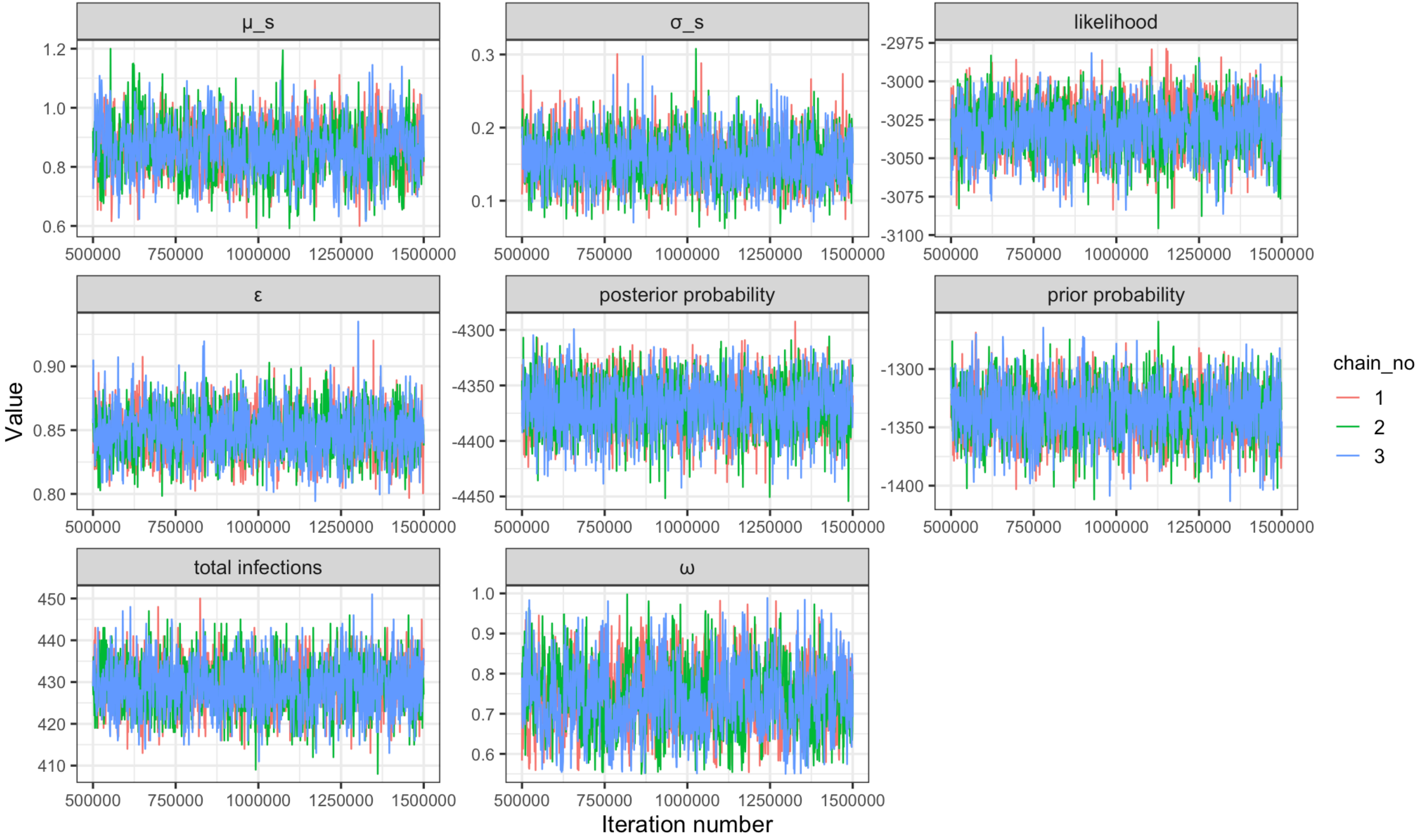

### S10 Fig

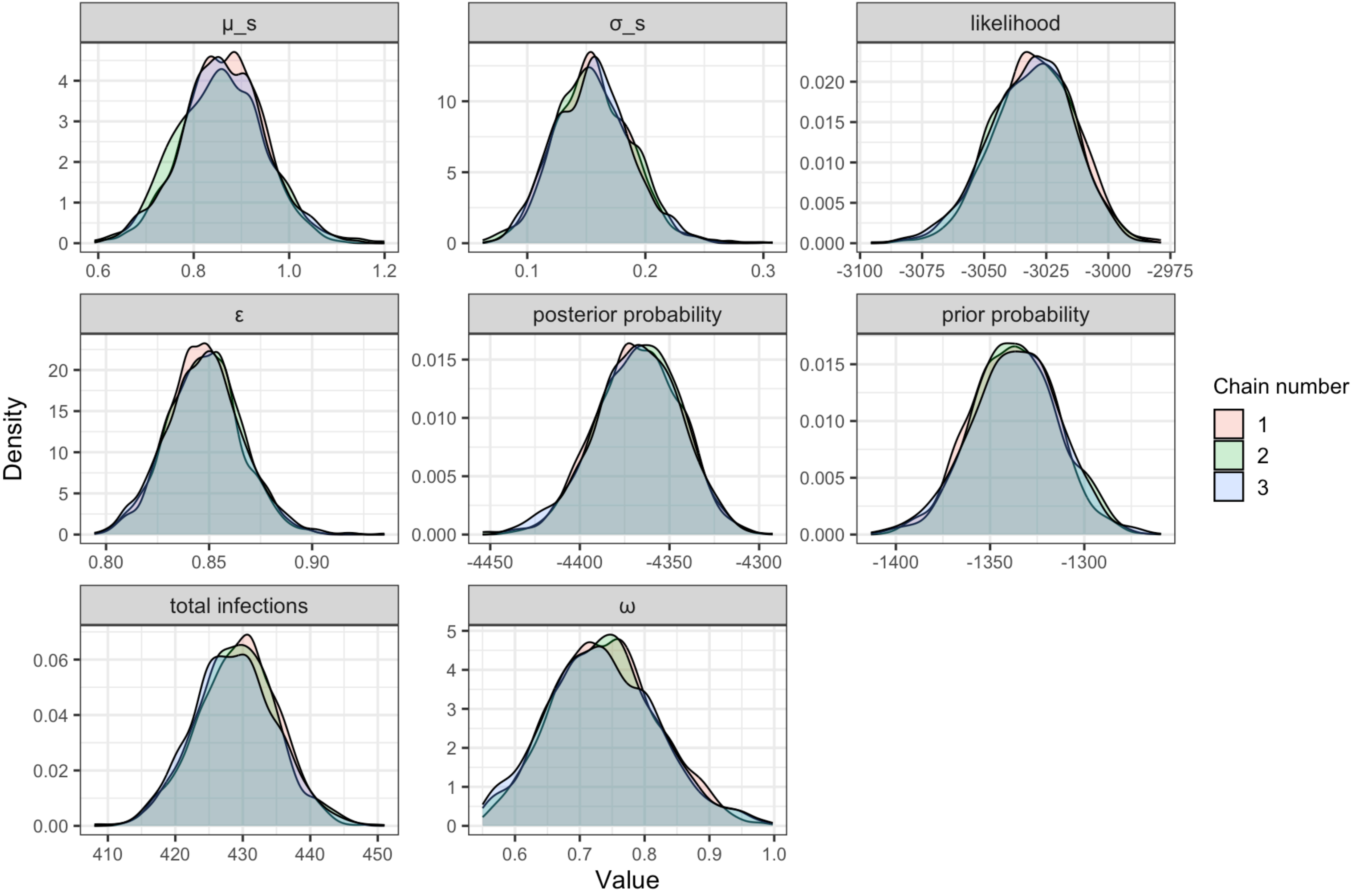

### S11 Fig

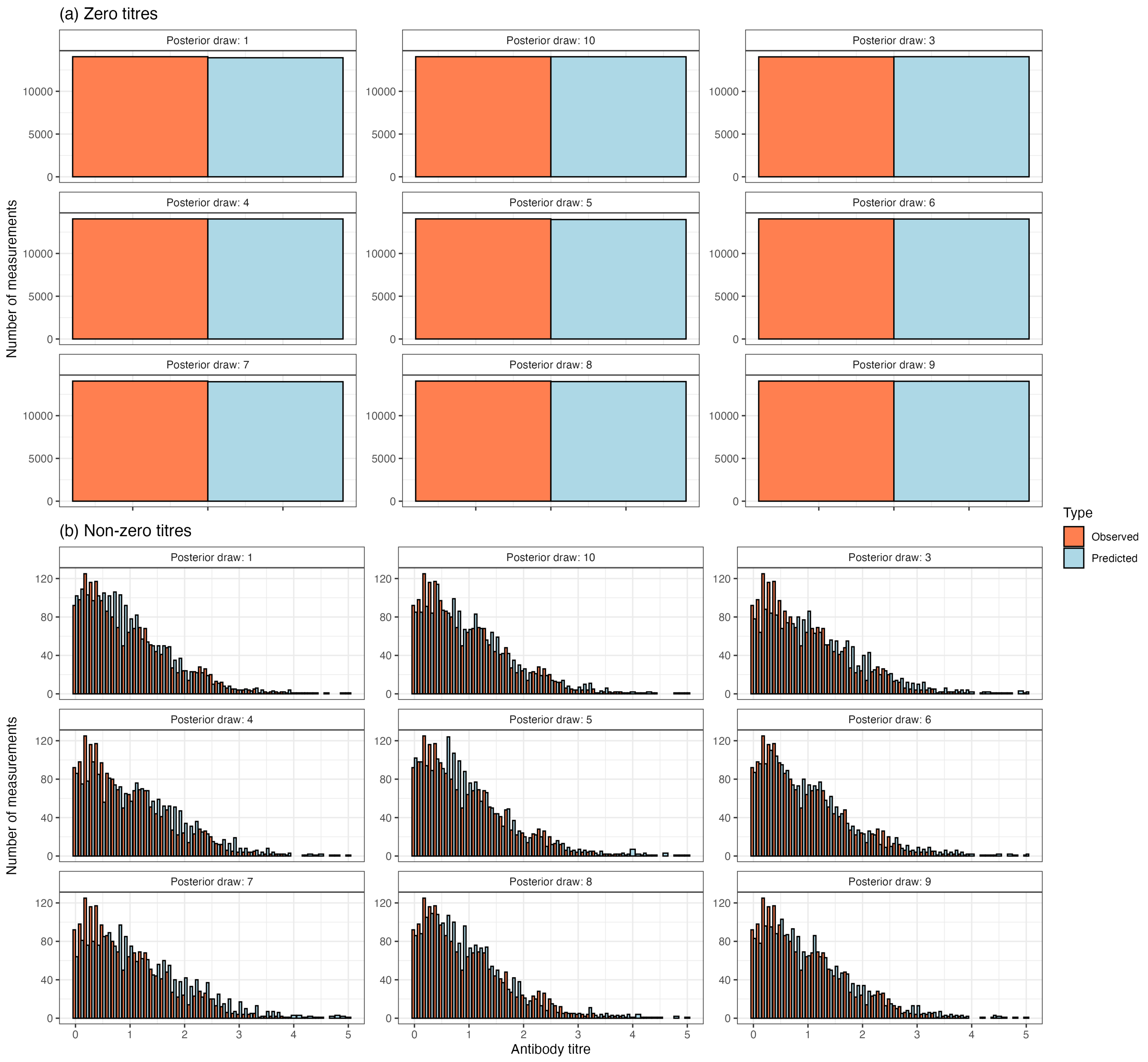

### S12 Fig

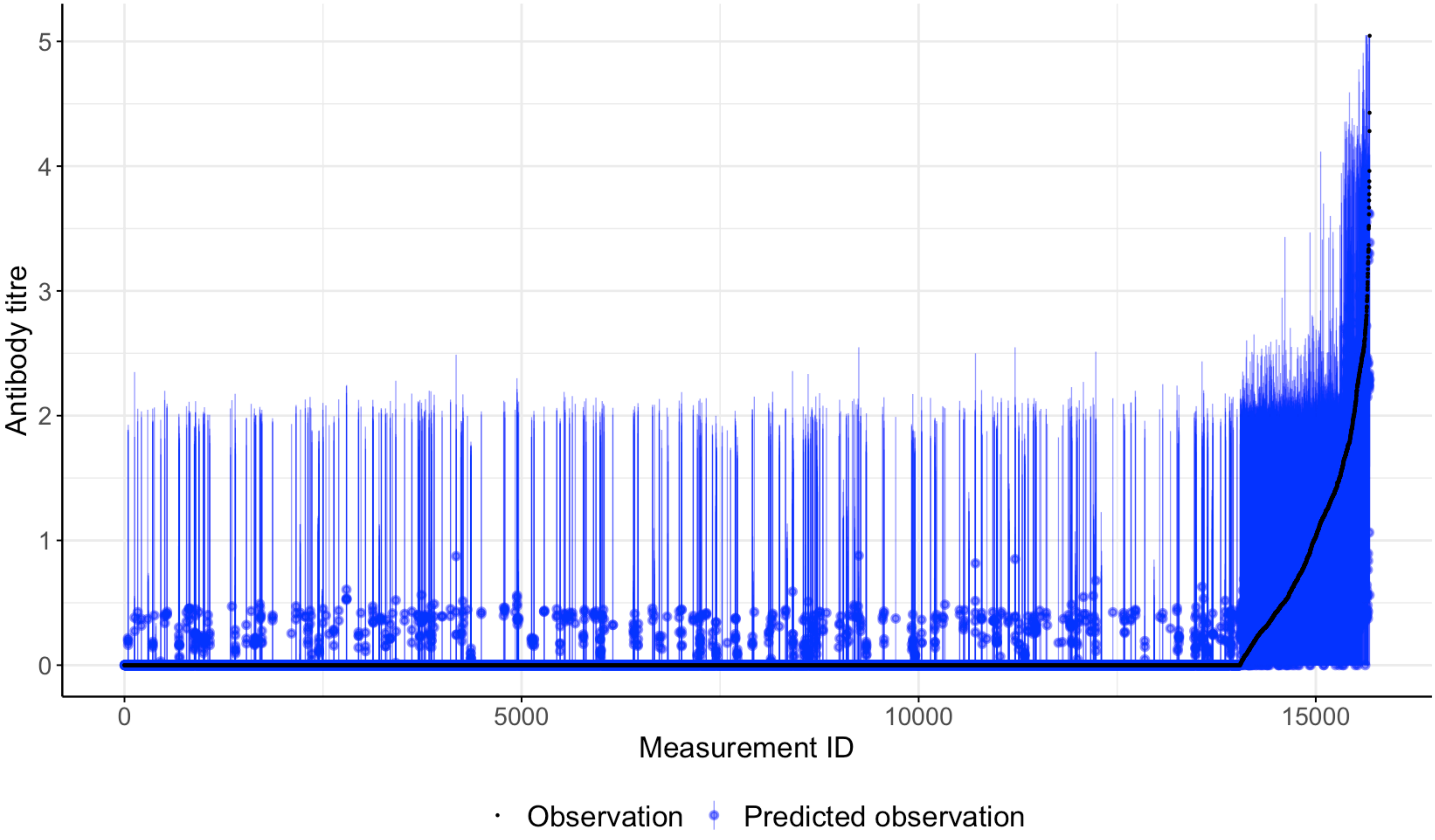

### S13 Fig

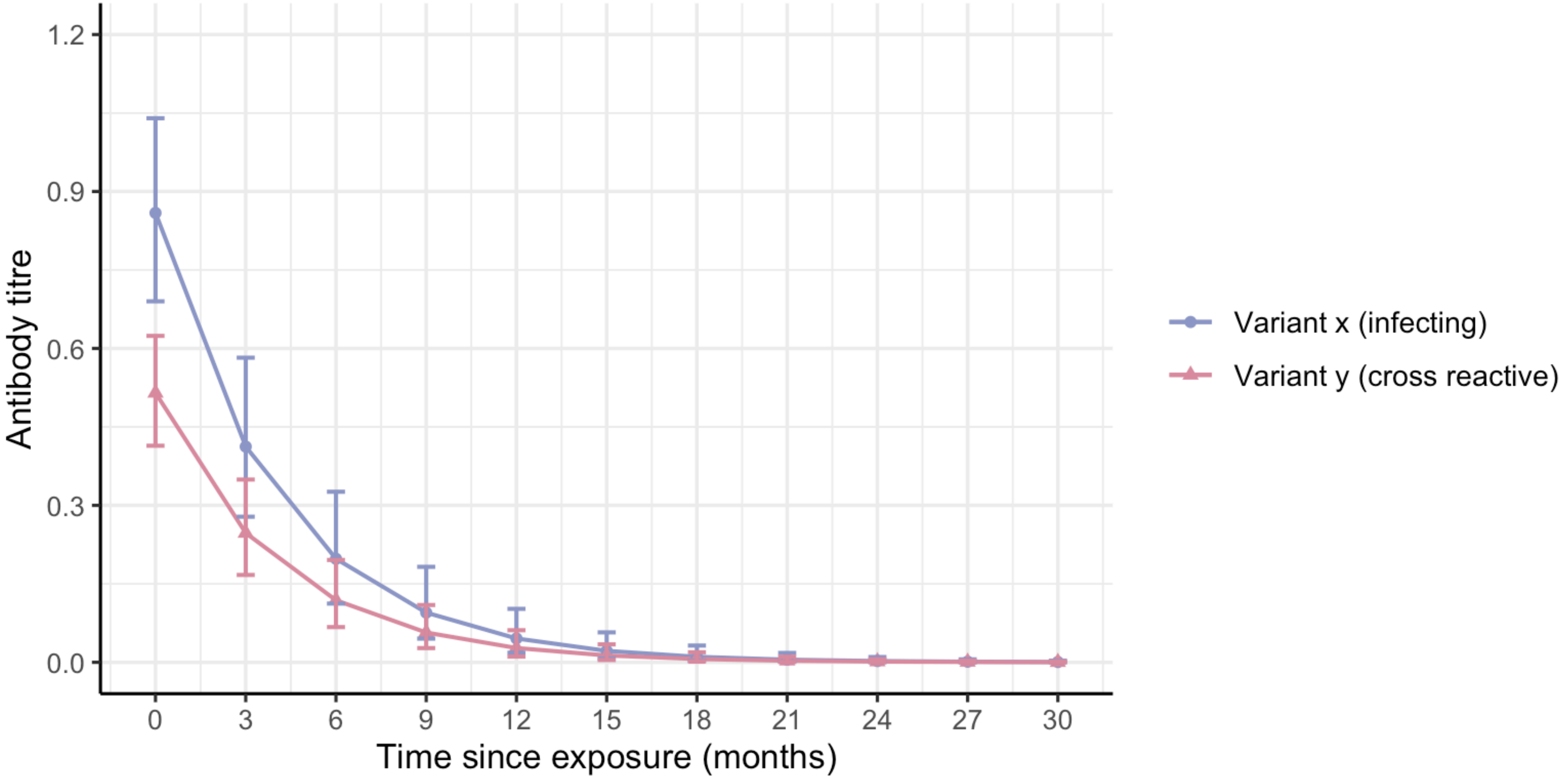

### S14 Fig

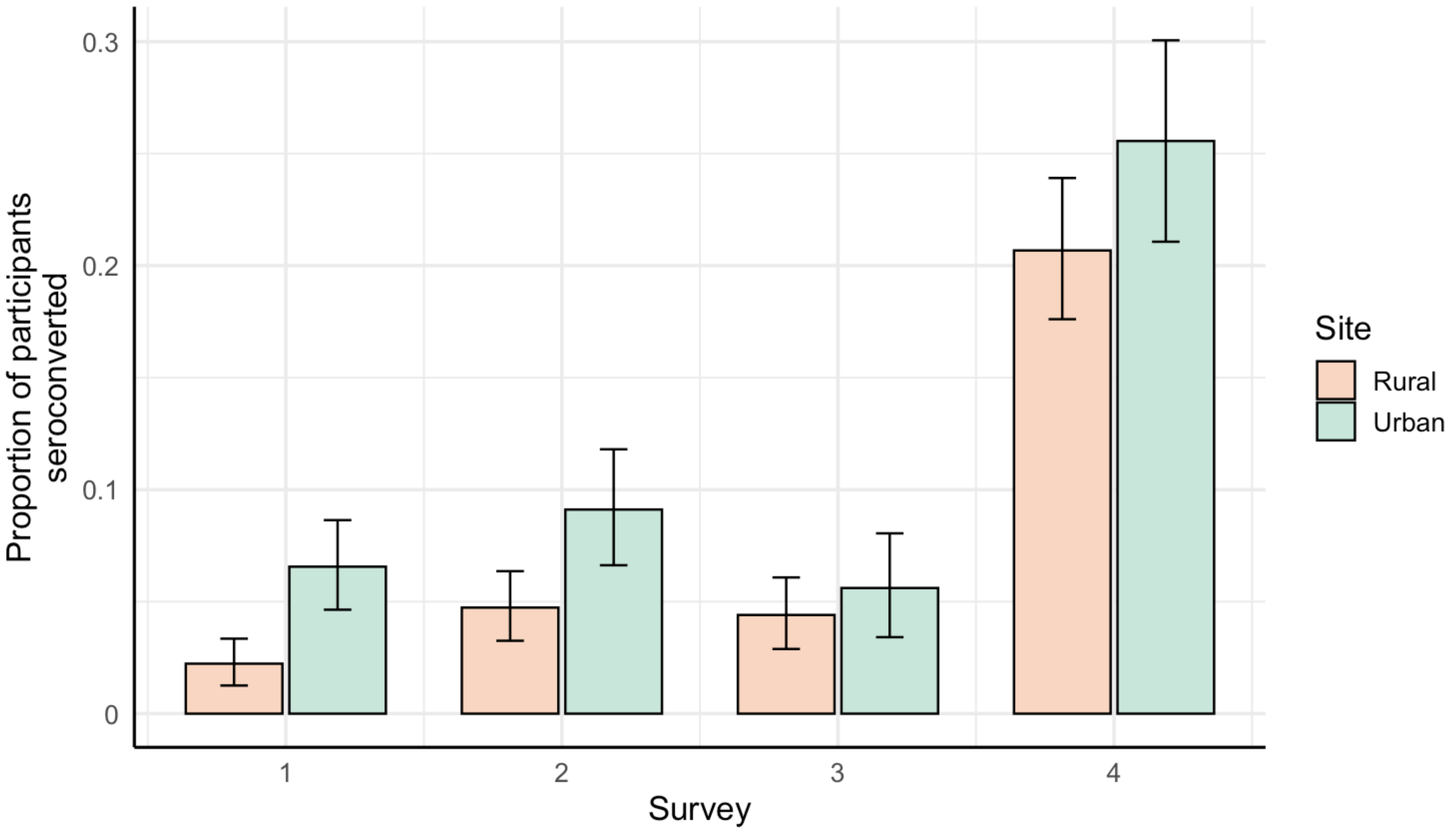

### S15 Fig

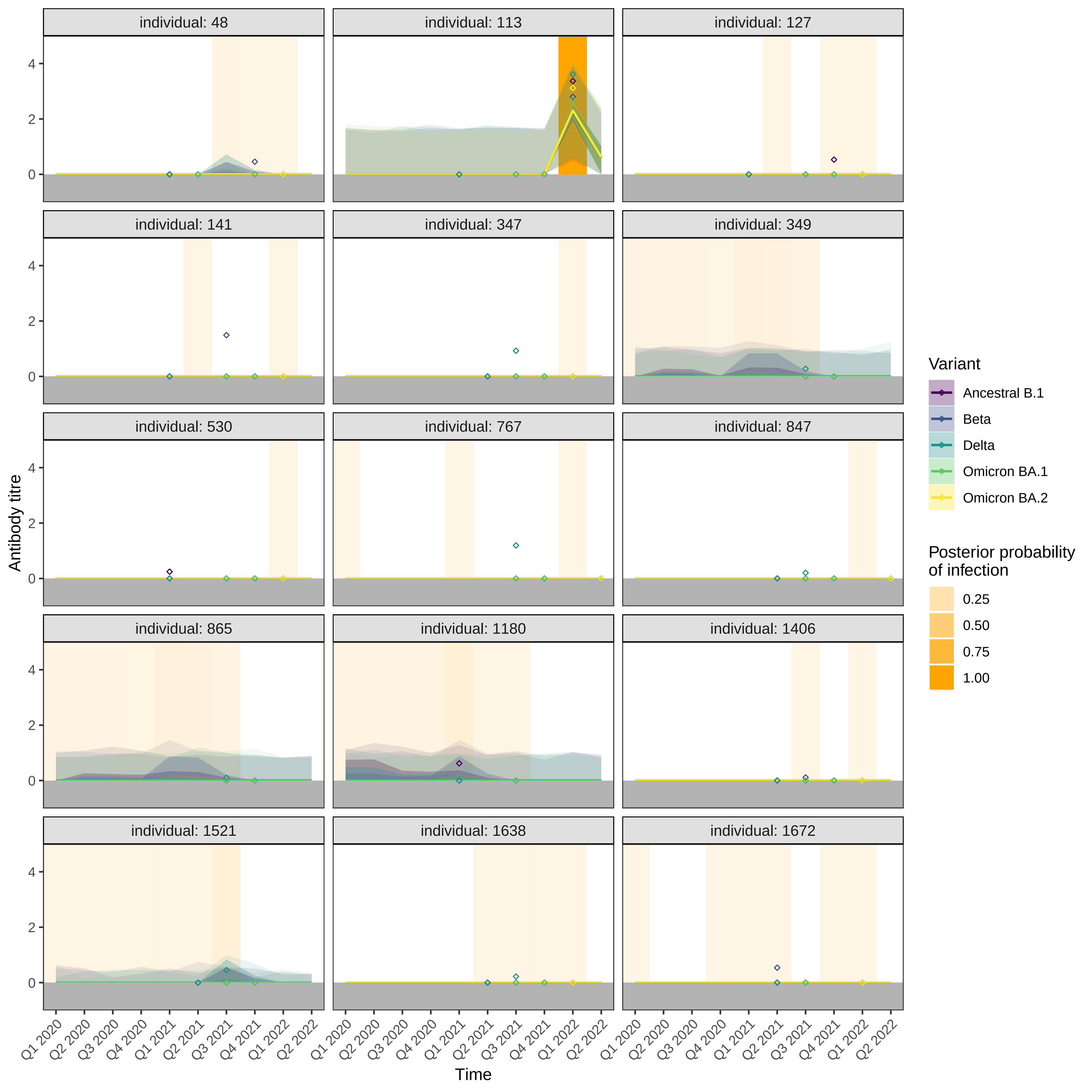

### S16 Fig

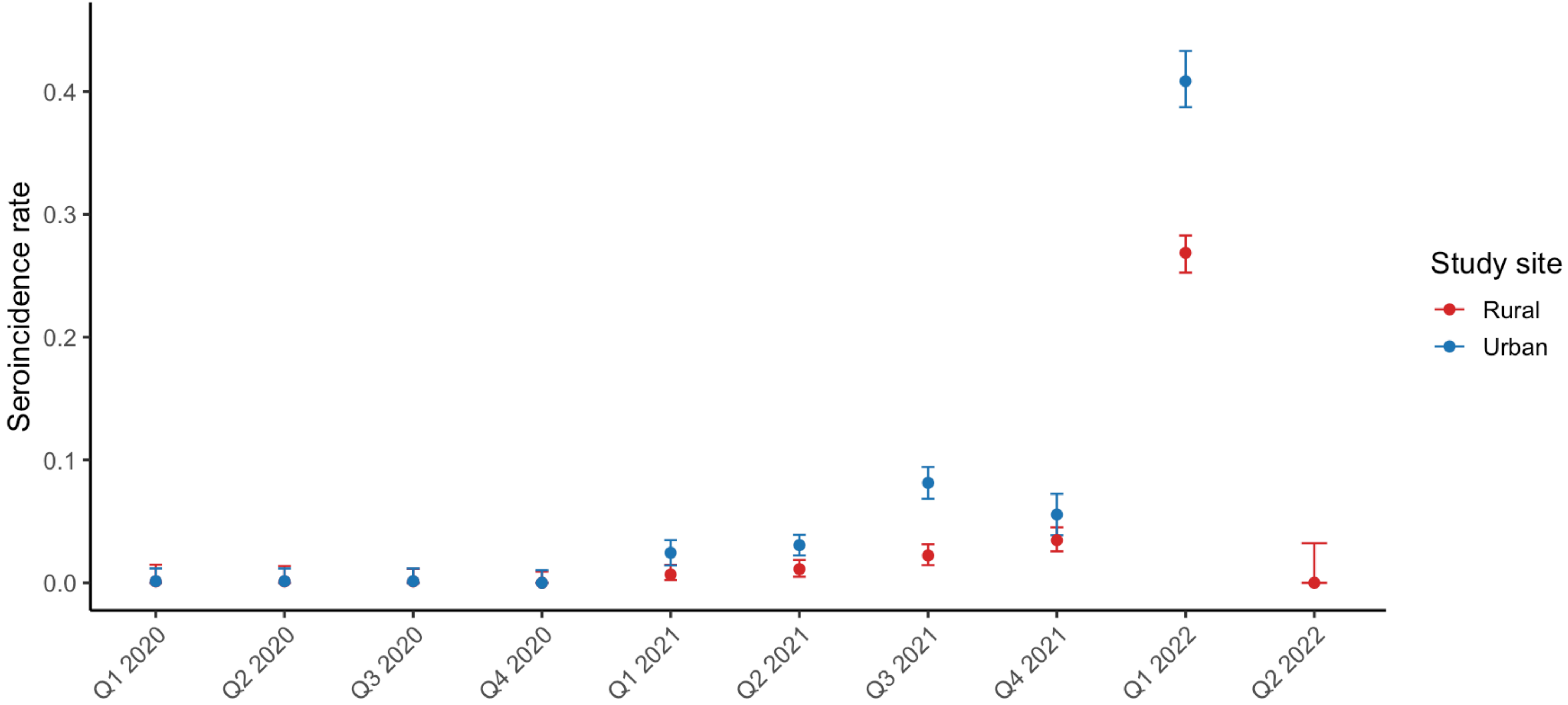
